# Oral rehydration salts and zinc for childhood diarrhoea in Bangladesh, 2019 and 2025: coverage, wealth inequality and reported source of care

**DOI:** 10.64898/2026.11.03.26366928

**Authors:** S. M. Ashraf Ul Hoque, Mohammad Khalid Hasan, Md Ashifur Rahman, Md Khairul Islam, Tanvir Ahmed, S M Raysul Haque, Md. Rabiul Islam, Kamran ul Baset

**Affiliations:** Department of Public Health, Independent University, Bangladesh (IUB); Department of Medicine, Dhaka Medical College Hospital

## Abstract

**Background:** Diarrhoea remains a major cause of morbidity and mortality among children under five. WHO and UNICEF recommend oral rehydration salts (ORS) alongside 10 to 14 days of zinc supplementation. However, national progress in combined treatment coverage and its socioeconomic and care-related inequalities remains insufficiently understood in Bangladesh. We assessed combined coverage, wealth inequality and differences by reported source of care.

**Methods:** In this repeated cross-sectional study, we analysed children under five with caregiver-reported diarrhoea in the 2019 (n=1,589) and 2025 (n=1,634) Multiple Indicator Cluster Surveys (MICS). We estimated combined coverage using design-based methods, wealth inequality using slope indices, and associations with source of care using pooled adjusted prevalence ratios (aPR). Demographic and Health Survey (DHS) estimates provided an external comparison.

**Results:** Combined ORS and zinc coverage increased from 35.0% (95% CI 32.3 to 37.7) in 2019 to 42.9% (40.1 to 45.8) in 2025. This was an increase of 7.9 points (4.0 to 11.8). In 2025, ORS and zinc coverage were 75.8% and 51.9%, respectively. The 2025 estimate was similar to DHS 2022 (difference −0.4 points, −7.0 to 6.3), whereas MICS 2019 was below DHS 2017-18. Unadjusted wealth inequality persisted in 2025 (slope index 13.0 percentage points, 3.4 to 22.6). Adjustment for sociodemographic characteristics attenuated the pooled slope index from 11.8 to 6.5 points (−1.7 to 14.6). Coverage was lower with pharmacy-only than public care (aPR 0.59, 0.53 to 0.66) and with no care reported (0.14, 0.11 to 0.18). Children with no care reported accounted for 43.7% of children without combined treatment in 2025.

**Conclusions:** Combined coverage increased within MICS, but fewer than half of children received both, and zinc remained the larger gap. Wealth inequality was partly shared with education and region. Children with no care reported formed the largest group without both treatments and need reaching outside facilities.

## Introduction

Diarrhoea remains a leading cause of death in children under five in low- and middle-income countries (LMICs) [1–3]. Most of these deaths follow dehydration, which oral rehydration salts (ORS) can prevent [4]. In children older than six months, zinc may shorten diarrhoea by about half a day, but trials show no clear benefit in younger infants [5]. The World Health Organization (WHO) and the United Nations Children’s Fund (UNICEF) have recommended since 2004 that every child with diarrhoea receive low-osmolarity ORS and zinc [6]. That statement gave a zinc course of 10 to 14 days, and WHO revised the zinc dose in 2024 [7].

In 2013, the Global Action Plan for Pneumonia and Diarrhoea (GAPPD) set a 2025 target of 90% access to appropriate case management [8]. It also set at least 80% coverage in every district [8]. Receiving both ORS and zinc is a narrower measure, so we use this target as context only. ORS coverage in LMICs reached only 44% by 2015 [9,10]. UNICEF estimates that about 22% of children with diarrhoea worldwide now receive both ORS and zinc [11].

Bangladesh sharply reduced diarrhoea deaths through work on ORS, zinc, hygiene and nutrition by public, non-governmental organisation (NGO) and private providers [12]. Diarrhoea deaths fell from 66.5 per 100,000 children in 2003 to 12.62 in 2019 [13]. From 2006, the Scaling Up Zinc for Young Children (SUZY) project scaled up zinc treatment nationally through public, private and informal providers [14]. Zinc use was at first higher in urban and richer families, but these gaps narrowed within two years [15]. The 2017-18 and 2022 Demographic and Health Surveys (DHS) found that about 50% of children with diarrhoea received zinc [13,16].

Three gaps remain. First, DHS combined coverage stayed near 43% in 2017-18 and in 2022 [16]. Yet many analyses track ORS and zinc separately [13], so little is known about which children miss combined treatment. Second, a recent study reported that zinc coverage in the 2022 DHS was within 5% across wealth quintiles [13]. It compared only the poorest and richest fifths, without a formal assessment of wealth-related inequality. Inequality indices based on wealth rank use data from every child and can show differences that such comparisons miss [17,18]. Third, little is known in Bangladesh about whether ORS and zinc use differs by source of care, the place where families seek advice or treatment. Earlier analyses of the Multiple Indicator Cluster Surveys (MICS) examined oral rehydration therapy and any care seeking, but not ORS and zinc by source of care [19,20]. Yet studies from several countries link source of care with how diarrhoea is treated [21], and poorer families more often use pharmacies and shops [22,23]. In Bangladesh, drug shops and village doctors, who are informal practitioners with short-course training and no medical degree, are the first place of care for many families [24,25]. They often dispense antibiotics for diarrhoea, and it is unclear how often they give zinc [26,27]. They also work under weak regulation and have strong incentives to sell medicines [28,29].

MICS 2025 was carried out in the GAPPD target year, six years after MICS 2019, and both asked the same questions on ORS and zinc [30,31]. Our aim was to find out how many children under five with diarrhoea in Bangladesh received both ORS and zinc, and which children missed out. We estimated coverage of ORS, zinc and both in 2019 and 2025. We then examined how combined coverage differed by household wealth, other characteristics and reported source of care.

## Materials and methods

### Data sources and study population

MICS are nationally representative household surveys conducted by the Bangladesh Bureau of Statistics (BBS) with UNICEF [30,31]. Each used a stratified two-stage cluster sample, with the urban and rural parts of each district as strata. The surveys selected 20 households in each of 3,220 clusters in 2019 and 3,149 clusters in 2025. Fieldwork ran from January to June 2019 and from February to June 2025. Trained interviewers used tablet-based computer-assisted personal interviewing with the standard MICS questionnaires, translated into Bangla and pre-tested. Supervisors monitored fieldwork daily, and one household per cluster was re-interviewed as a quality check. Mothers or caregivers answered the questionnaire for children under five. We used the 2006 and 2012-13 rounds for ORS context only [32,33], and S1 Fig shows their estimates.

The caregiver was asked: “In the last two weeks, has (name) had diarrhoea?” The measure is therefore caregiver recognition of diarrhoea, with no clinical definition. Of 23,101 children with a valid answer in 2019 and 23,365 in 2025, 1,589 and 1,634 had diarrhoea and formed the study population. These include 2 and 8 children from partly completed interviews, given zero weight. We followed the Strengthening the Reporting of Observational Studies in Epidemiology (STROBE) guideline [34].

### Coverage of ORS, zinc and both

The outcomes were receipt of ORS, zinc and both during the episode. Both meant reported receipt of any ORS and any zinc, not simultaneous use or a complete course. A child counted as receiving ORS if the caregiver reported fluid from an ORS packet or a rice-based ORS packet, as in MICS indicator TC.13a [30]. Home-made sugar-salt solution, coconut water and rice water were not counted. Caregivers were asked directly whether the child was given zinc tablets or syrup. The surveys did not record ORS osmolarity or the dose and duration of zinc (S1 Table). Following MICS rules [30], the 12 “don’t know” answers (S1 Table) were counted as not given. We analysed zinc only for 2019 and 2025, because MICS 2012-13 recorded zinc only from unprompted recall. The 2025 report gives 41.6% for combined treatment, a figure we could reproduce only by counting packet ORS alone. We counted any ORS in both surveys, to match the ORS indicator in both reports and the DHS. This harmonised definition gave 42.9% in 2025 (S1 Table).

### Wealth and source of care

We ranked children with diarrhoea by the MICS wealth score of each survey, an index built by principal component analysis of household assets, dwelling, water and sanitation [30,31]. Each child received a weighted fractional rank from 0 (poorest) to 1 (richest; S5 Table).

Caregivers were asked whether advice or treatment was sought for the diarrhoea, and from which places. We placed each child in the first of five groups that applied, in the order below. Public care meant any government hospital, health centre, community clinic, community health worker or outreach clinic. Private facility or physician meant a private or NGO hospital, clinic, physician or health worker. Pharmacy only meant a private pharmacy and no other medical source, and the outlets behind this code are not observed, so it may include shops that sell medicines. Some pharmacy-only children also named an informal source (18 in 2019 and 13 in 2025; S5 Table). Traditional or other informal meant a traditional practitioner, relative, shop or other source. The fifth group was no care sought. Of care seekers, 5.7% in 2019 and 2.6% in 2025 named more than one type of source (S1 and S5 Tables).

The lists had no code for village doctors, whose care may have been recorded under private physicians, pharmacies or traditional practitioners. The lists were the same in both surveys, except that 2025 split NGO care into three codes (S1 Table). Contrasts between groups are therefore not contrasts between provider types. In 2019 only, caregivers were asked where they obtained ORS and zinc (questionnaire items CA9 and CA11).

### Statistical analysis

After reproducing the published national estimates, we estimated coverage by sex, age, urban or rural area, division, mother’s education and wealth quintile. Age had six groups, with infants split into those under six months and those aged 6 to 11 months. Regression models used 0 to 11 months as one group, as in the survey reports. Change between surveys was the difference between the two estimates, with the samples treated as independent.

We used DHS 2017-18 and 2022 estimates as an external comparison, not as a study aim [16]. The DHS differs in its denominator (living children born in the past five years), its second ORS item and its interview months. We re-estimated DHS confidence intervals from the data files, and point estimates matched published values within 0.1 points (S1 Table). We compared levels and changes between programmes with z tests.

The slope index of inequality (SII) is the predicted difference in coverage, in percentage points, between the richest and poorest ends of the wealth distribution [17]. We estimated it from a survey-weighted Gaussian model with an identity link. In adjusted models, the SII is the rank coefficient conditional on the other covariates. We tested change in the SII with a rank-by-year interaction and linearity with squared and cubic rank terms. To compare the zinc and ORS gradients, we modelled the difference between the two outcomes for each child against wealth rank.

To see how much of the wealth gradient was shared with other characteristics and source of care, we added each set of covariates to the SII model. Each pooled SII model included survey year, so it assumed one rank slope with a separate intercept for each survey. Each reduction was the paired difference between two models within every replicate (S3 Table). This statistical attenuation is not mediation, because source of care was measured at the same time as treatment. We also estimated by wealth the share of children who received care from a public or private facility or physician.

We estimated adjusted prevalence ratios (aPR) with survey-weighted Poisson regression and a log link, because combined treatment was common [35,36]. Model 1 pooled both surveys and included survey year, wealth quintile, mother’s education, area, division, child age group and sex. These covariates were chosen a priori from earlier survey analyses [19,20,37,38]. Model 2 added source of care. We tested a survey-by-source interaction and report year-specific aPRs for source of care. A survey-weighted linear model tested whether gains differed between sources on the additive scale. As a descriptive addition, a two-fold Kitagawa decomposition split the 2019 to 2025 change into change within sources and shifts between sources [39]. Exploratory survey-by-characteristic interactions were added one at a time, and subgroup comparisons were descriptive and not corrected for multiple testing (S5 Table).

All estimates used child sampling weights and the clusters and strata of the full child sample, with children with diarrhoea analysed as a subpopulation. In pooled analyses, strata and clusters were kept distinct by survey (259 strata and 6,362 clusters in the full child sample). Children with diarrhoea came from 2,268 clusters in 251 strata, which gave 2,017 design degrees of freedom. Weights averaged 1.00 in each survey and were pooled without rescaling, so each survey contributes in proportion to its sample. Single-cluster strata were centred at the grand mean. Standard errors came from Taylor series linearisation, and Wald tests used design-based degrees of freedom. Intervals for the attenuation and the decomposition came from 1,000 Rao-Wu subbootstrap replicates. These resampled clusters within strata of each survey, kept wealth ranks fixed and used normal intervals. Estimates are given with 95% confidence intervals (CIs) and two-sided p values.

Data were complete except for one child with missing wealth and age, so the pooled models used 3,222 of 3,223 children. We did 13 sensitivity analyses, each testing one assumption (S5 Table). Five tests of the change used packet ORS only, “don’t know” answers as missing, ages 6 to 59 months, adjustment for interview month, and February to May only. Other tests varied the wealth ranking, the order of assigning source of care, illness proxies, infant age groups and the regression model. Because village doctor care may fall under any of these codes, we combined the private physician, pharmacy and traditional groups and compared their share between surveys.

Analyses were done in Stata version 17 (StataCorp, College Station, Texas, USA) and R version 4.6.1 with the survey package version 4.5 [40].

### Ethics

MICS 2019 was approved by a technical committee of the Government of Bangladesh led by BBS [31]. MICS 2025 was approved by the Ethical Review Committee of the International Centre for Diarrhoeal Disease Research, Bangladesh (icddr,b; PR-24113) [30]. Both reports state that verbal informed consent was obtained. The 2017-18 and 2022 DHS protocols were approved by the ICF Institutional Review Board and the Bangladesh Medical Research Council [41,42]. Our analysis used anonymised, publicly available data and did not need further ethical approval.

## Results

The weighted prevalence of diarrhoea was 6.9% (95% CI 6.5 to 7.3) in 2019 and 7.1% (6.7 to 7.5) in 2025 (S1 Table). Response rates for the under-five questionnaire were 93.6% in 2019 and 94.6% in 2025 [30,31]. Children with diarrhoea in 2025 had better-educated mothers and more often lived in urban areas (Table 1). Our estimates matched the survey reports to one decimal place for all indicators defined the same way (S1 Table).

**Table 1.** Characteristics of children under five with diarrhoea in the past two weeks, MICS 2019 and 2025. Values are weighted percentages, with unweighted and weighted numbers in brackets.

| Characteristic | Group | 2019, % (n; weighted n) | 2025, % (n; weighted n) |
|---|---|---|---|
| Sex | Male | 53.9 (852; 860) | 52.9 (873; 873) |
|  | Female | 46.1 (737; 736) | 47.1 (761; 778) |
| Age (months) | 0-5 | 10.1 (155; 161) | 8.4 (134; 139) |
|  | 6-11 | 16.3 (254; 260) | 17.7 (285; 292) |
|  | 12-23 | 28.1 (459; 448) | 30.4 (483; 501) |
|  | 24-35 | 20.4 (330; 326) | 19.7 (337; 325) |
|  | 36-47 | 15.5 (232; 247) | 13.2 (222; 218) |
|  | 48-59 | 9.6 (158; 154) | 10.7 (173; 176) |
| Area | Urban | 21.4 (299; 342) | 27.0 (409; 446) |
|  | Rural | 78.6 (1,290; 1,255) | 73.0 (1,225; 1,205) |
| Division | Barishal | 11.6 (258; 185) | 6.9 (171; 114) |
|  | Chattogram | 23.8 (385; 380) | 32.8 (491; 541) |
|  | Dhaka | 19.5 (249; 311) | 20.6 (290; 341) |
|  | Khulna | 9.7 (204; 155) | 8.1 (168; 134) |
|  | Mymensingh | 9.6 (113; 153) | 6.8 (87; 112) |
|  | Rajshahi | 11.4 (152; 182) | 10.7 (155; 177) |
|  | Rangpur | 7.0 (116; 112) | 8.1 (164; 134) |
|  | Sylhet | 7.4 (112; 119) | 5.9 (108; 97) |
| Mother’s education | No education | 12.5 (201; 199) | 5.8 (93; 96) |
|  | Primary | 25.2 (425; 402) | 20.0 (332; 330) |
|  | Secondary | 48.8 (758; 779) | 53.9 (879; 891) |
|  | Higher secondary or more | 13.6 (205; 217) | 20.2 (330; 334) |
| Wealth quintile | Lowest (Quintile 1 / Poorest) | 26.4 (489; 421) | 21.8 (406; 361) |
|  | Second (Quintile 2 / Poorer) | 23.2 (379; 371) | 20.3 (342; 336) |
|  | Middle (Quintile 3) | 16.4 (257; 262) | 18.9 (332; 311) |
|  | Fourth (Quintile 4 / Richer) | 17.6 (267; 281) | 20.5 (297; 339) |
|  | Highest (Quintile 5 / Richest) | 16.4 (196; 262) | 18.4 (257; 304) |
| Source of care | Public health care | 12.6 (201; 201) | 10.0 (172; 165) |
|  | Private facility or physician | 16.9 (270; 270) | 12.2 (202; 201) |
|  | Pharmacy only | 25.9 (368; 413) | 35.0 (592; 579) |
|  | Traditional or other informal | 17.2 (303; 274) | 14.2 (213; 235) |
|  | No care sought | 27.5 (447; 438) | 28.5 (455; 471) |

In each cell, the first number in brackets is the number of children interviewed (1,589 in 2019 and 1,634 in 2025). The second is the weighted number, the sum of sampling weights, which totals 1,596 in 2019 and 1,651 in 2025. Percentages use the weighted numbers. One child in 2019 had no recorded age or wealth and a sampling weight of zero. The age and wealth rows for 2019 therefore sum to 1,588 children. Wealth quintiles are national household quintiles, so their shares among children with diarrhoea are not 20%. Rounded weighted numbers may not sum exactly to the totals.

### Coverage of ORS, zinc and both

ORS coverage was 72.4% (95% CI 69.9 to 74.8) in 2019 and 75.8% (73.4 to 78.2) in 2025 (Table 2, S2 Table). The rise was 3.4 points (0.0 to 6.9; p=0.048). Zinc coverage rose from 43.6% (40.7 to 46.4) to 51.9% (49.0 to 54.8; S2 Table). Combined coverage rose from 35.0% (32.3 to 37.7) to 42.9% (40.1 to 45.8), an increase of 7.9 percentage points (4.0 to 11.8; Table 2). Among children given ORS, the share also given zinc was 48.4% in 2019 and 56.6% in 2025 (S4 Table). In 2025, 32.9% received ORS without zinc and 15.2% received neither (S4 Table).

**Table 2.**
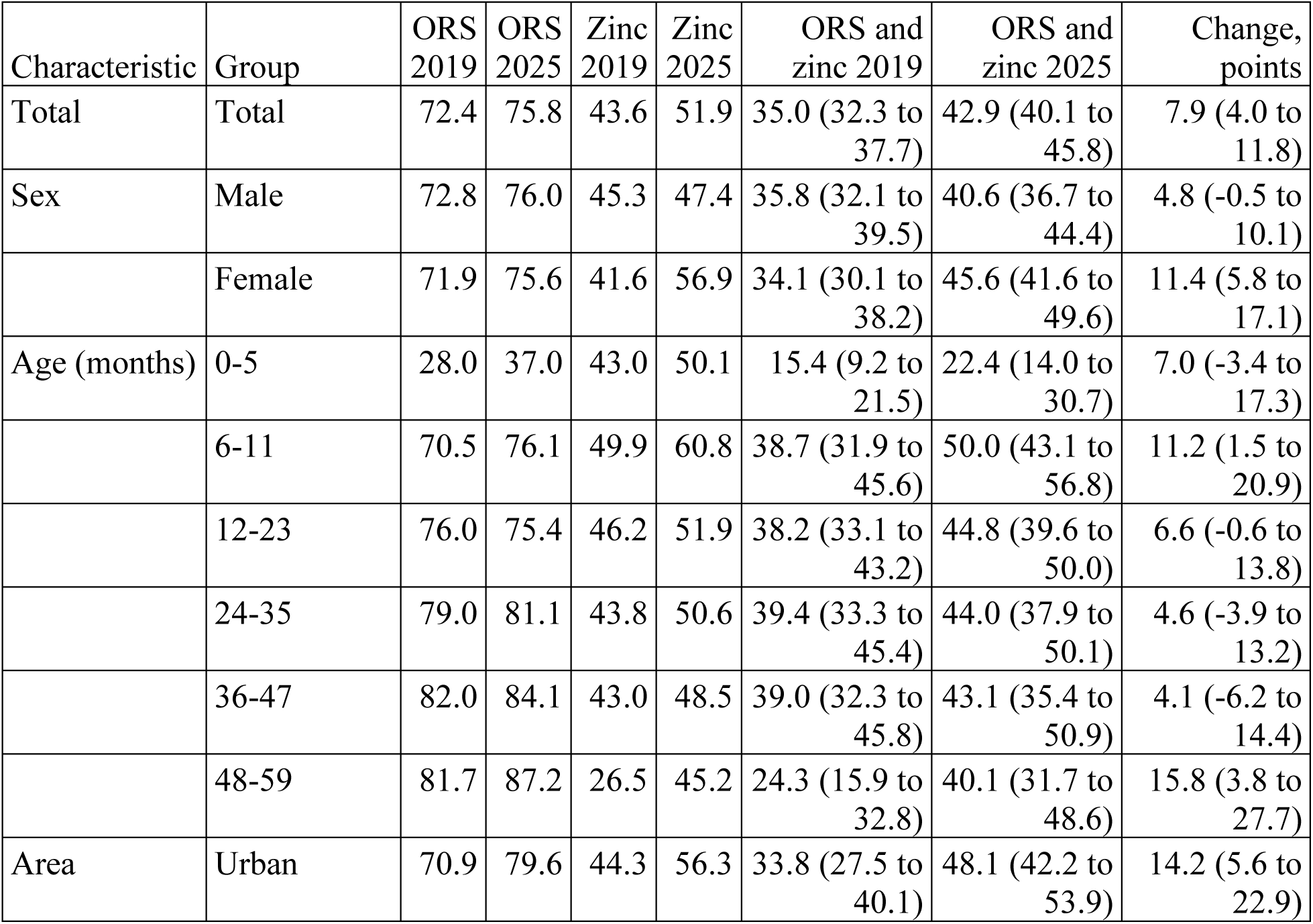
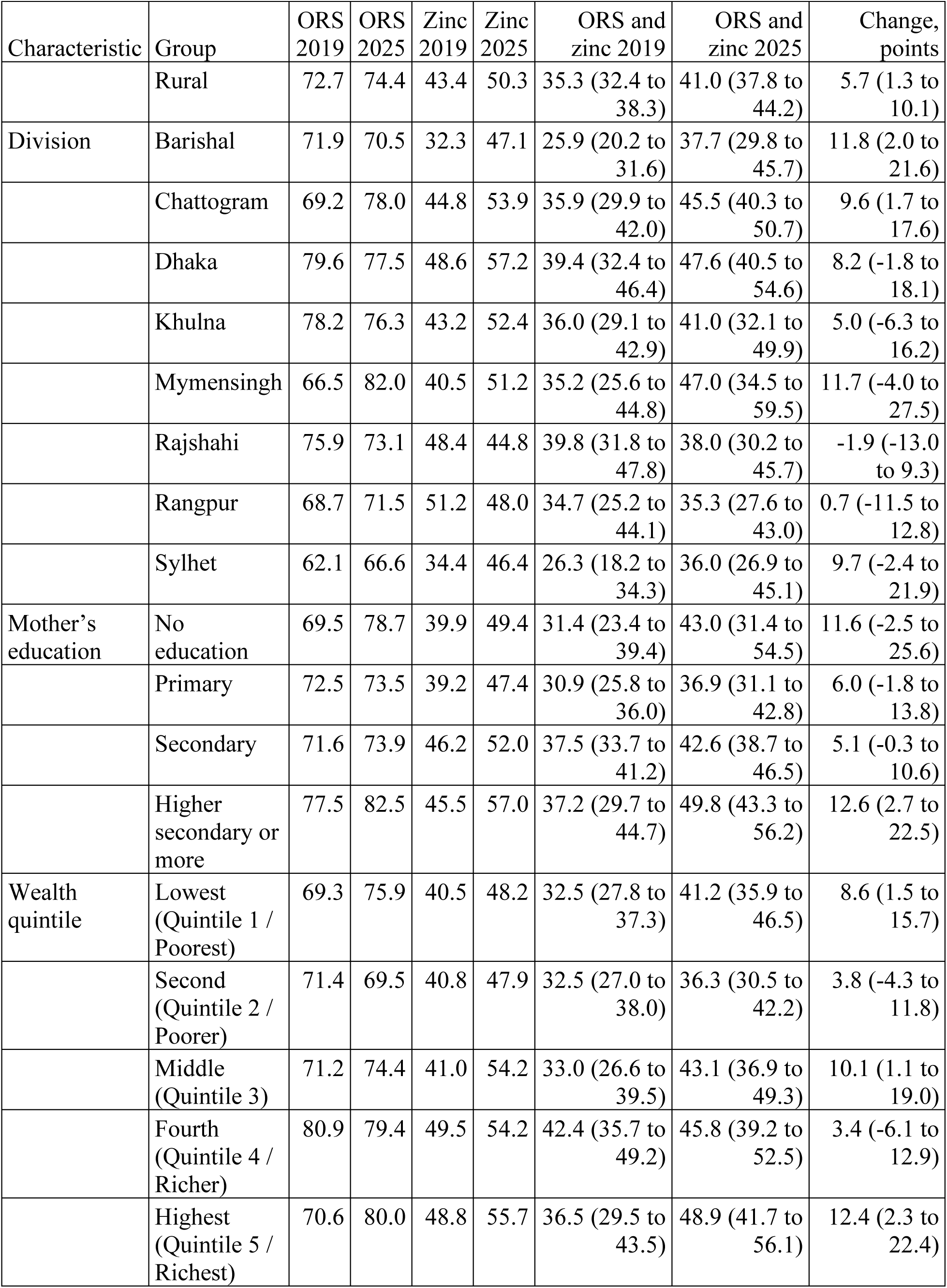
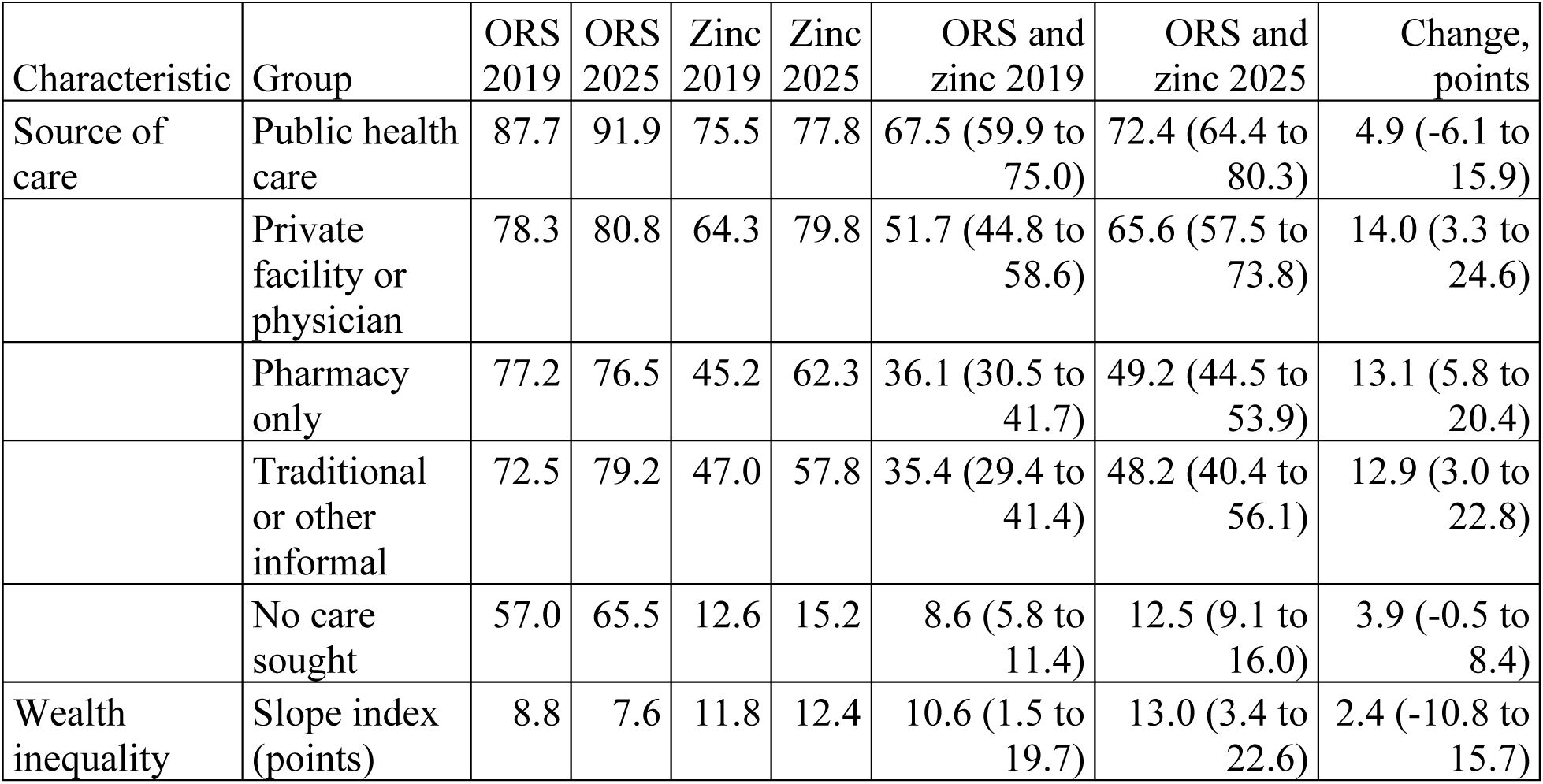
Coverage of ORS, zinc and combined treatment and wealth-related inequality among children with diarrhoea, MICS 2019 and 2025. Values are weighted percentages. The 95% confidence intervals (CIs) are shown for combined treatment and its change. S2 Table gives CIs for ORS and zinc.

| Characteristic | Group | ORS 2019 | ORS 2025 | Zinc 2019 | Zinc 2025 | ORS and zinc 2019 | ORS and zinc 2025 | Change, points |
|---|---|---|---|---|---|---|---|---|
| Total | Total | 72.4 | 75.8 | 43.6 | 51.9 | 35.0 (32.3 to 37.7) | 42.9 (40.1 to 45.8) | 7.9 (4.0 to 11.8) |
| Sex | Male | 72.8 | 76.0 | 45.3 | 47.4 | 35.8 (32.1 to 39.5) | 40.6 (36.7 to 44.4) | 4.8 (-0.5 to 10.1) |
|  | Female | 71.9 | 75.6 | 41.6 | 56.9 | 34.1 (30.1 to 38.2) | 45.6 (41.6 to 49.6) | 11.4 (5.8 to 17.1) |
| Age (months) | 0-5 | 28.0 | 37.0 | 43.0 | 50.1 | 15.4 (9.2 to 21.5) | 22.4 (14.0 to 30.7) | 7.0 (-3.4 to 17.3) |
|  | 6-11 | 70.5 | 76.1 | 49.9 | 60.8 | 38.7 (31.9 to 45.6) | 50.0 (43.1 to 56.8) | 11.2 (1.5 to 20.9) |
|  | 12-23 | 76.0 | 75.4 | 46.2 | 51.9 | 38.2 (33.1 to 43.2) | 44.8 (39.6 to 50.0) | 6.6 (-0.6 to 13.8) |
|  | 24-35 | 79.0 | 81.1 | 43.8 | 50.6 | 39.4 (33.3 to 45.4) | 44.0 (37.9 to 50.1) | 4.6 (-3.9 to 13.2) |
|  | 36-47 | 82.0 | 84.1 | 43.0 | 48.5 | 39.0 (32.3 to 45.8) | 43.1 (35.4 to 50.9) | 4.1 (-6.2 to 14.4) |
|  | 48-59 | 81.7 | 87.2 | 26.5 | 45.2 | 24.3 (15.9 to 32.8) | 40.1 (31.7 to 48.6) | 15.8 (3.8 to 27.7) |
| Area | Urban | 70.9 | 79.6 | 44.3 | 56.3 | 33.8 (27.5 to 40.1) | 48.1 (42.2 to 53.9) | 14.2 (5.6 to 22.9) |
|  | Rural | 72.7 | 74.4 | 43.4 | 50.3 | 35.3 (32.4 to 38.3) | 41.0 (37.8 to 44.2) | 5.7 (1.3 to 10.1) |
| Division | Barishal | 71.9 | 70.5 | 32.3 | 47.1 | 25.9 (20.2 to 31.6) | 37.7 (29.8 to 45.7) | 11.8 (2.0 to 21.6) |
|  | Chattogram | 69.2 | 78.0 | 44.8 | 53.9 | 35.9 (29.9 to 42.0) | 45.5 (40.3 to 50.7) | 9.6 (1.7 to 17.6) |
|  | Dhaka | 79.6 | 77.5 | 48.6 | 57.2 | 39.4 (32.4 to 46.4) | 47.6 (40.5 to 54.6) | 8.2 (-1.8 to 18.1) |
|  | Khulna | 78.2 | 76.3 | 43.2 | 52.4 | 36.0 (29.1 to 42.9) | 41.0 (32.1 to 49.9) | 5.0 (-6.3 to 16.2) |
|  | Mymensingh | 66.5 | 82.0 | 40.5 | 51.2 | 35.2 (25.6 to 44.8) | 47.0 (34.5 to 59.5) | 11.7 (-4.0 to 27.5) |
|  | Rajshahi | 75.9 | 73.1 | 48.4 | 44.8 | 39.8 (31.8 to 47.8) | 38.0 (30.2 to 45.7) | -1.9 (-13.0 to 9.3) |
|  | Rangpur | 68.7 | 71.5 | 51.2 | 48.0 | 34.7 (25.2 to 44.1) | 35.3 (27.6 to 43.0) | 0.7 (-11.5 to 12.8) |
|  | Sylhet | 62.1 | 66.6 | 34.4 | 46.4 | 26.3 (18.2 to 34.3) | 36.0 (26.9 to 45.1) | 9.7 (-2.4 to 21.9) |
| Mother’s education | No education | 69.5 | 78.7 | 39.9 | 49.4 | 31.4 (23.4 to 39.4) | 43.0 (31.4 to 54.5) | 11.6 (-2.5 to 25.6) |
|  | Primary | 72.5 | 73.5 | 39.2 | 47.4 | 30.9 (25.8 to 36.0) | 36.9 (31.1 to 42.8) | 6.0 (-1.8 to 13.8) |
|  | Secondary | 71.6 | 73.9 | 46.2 | 52.0 | 37.5 (33.7 to 41.2) | 42.6 (38.7 to 46.5) | 5.1 (-0.3 to 10.6) |
|  | Higher secondary or more | 77.5 | 82.5 | 45.5 | 57.0 | 37.2 (29.7 to 44.7) | 49.8 (43.3 to 56.2) | 12.6 (2.7 to 22.5) |
| Wealth quintile | Lowest (Quintile 1 / Poorest) | 69.3 | 75.9 | 40.5 | 48.2 | 32.5 (27.8 to 37.3) | 41.2 (35.9 to 46.5) | 8.6 (1.5 to 15.7) |
|  | Second (Quintile 2 / Poorer) | 71.4 | 69.5 | 40.8 | 47.9 | 32.5 (27.0 to 38.0) | 36.3 (30.5 to 42.2) | 3.8 (-4.3 to 11.8) |
|  | Middle (Quintile 3) | 71.2 | 74.4 | 41.0 | 54.2 | 33.0 (26.6 to 39.5) | 43.1 (36.9 to 49.3) | 10.1 (1.1 to 19.0) |
|  | Fourth (Quintile 4 / Richer) | 80.9 | 79.4 | 49.5 | 54.2 | 42.4 (35.7 to 49.2) | 45.8 (39.2 to 52.5) | 3.4 (-6.1 to 12.9) |
|  | Highest (Quintile 5 / Richest) | 70.6 | 80.0 | 48.8 | 55.7 | 36.5 (29.5 to 43.5) | 48.9 (41.7 to 56.1) | 12.4 (2.3 to 22.4) |
| Source of care | Public health care | 87.7 | 91.9 | 75.5 | 77.8 | 67.5 (59.9 to 75.0) | 72.4 (64.4 to 80.3) | 4.9 (-6.1 to 15.9) |
|  | Private facility or physician | 78.3 | 80.8 | 64.3 | 79.8 | 51.7 (44.8 to 58.6) | 65.6 (57.5 to 73.8) | 14.0 (3.3 to 24.6) |
|  | Pharmacy only | 77.2 | 76.5 | 45.2 | 62.3 | 36.1 (30.5 to 41.7) | 49.2 (44.5 to 53.9) | 13.1 (5.8 to 20.4) |
|  | Traditional or other informal | 72.5 | 79.2 | 47.0 | 57.8 | 35.4 (29.4 to 41.4) | 48.2 (40.4 to 56.1) | 12.9 (3.0 to 22.8) |
|  | No care sought | 57.0 | 65.5 | 12.6 | 15.2 | 8.6 (5.8 to 11.4) | 12.5 (9.1 to 16.0) | 3.9 (-0.5 to 8.4) |
| Wealth inequality | Slope index (points) | 8.8 | 7.6 | 11.8 | 12.4 | 10.6 (1.5 to 19.7) | 13.0 (3.4 to 22.6) | 2.4 (-10.8 to 15.7) |

Each value is the percentage of children with diarrhoea in the group who received the treatment, weighted with the survey sampling weights. Table 1 gives the number of children in each group. Change is the 2025 value minus the 2019 value, in percentage points. The slope index is the predicted difference in coverage between the richest and poorest ends of the wealth distribution, from a linear model on fractional wealth rank. Mymensingh (n=87 in 2025) and other small groups have wide intervals. The test of different gains between sources on the additive scale gave p=0.10.

Infants under six months had the lowest ORS coverage, at 28.0% (95% CI 20.6 to 35.5) in 2019 and 37.0% (27.6 to 46.3) in 2025 (Table 2). Their zinc coverage (43.0% and 50.1%) was similar to that of older children, so their combined coverage was 15.4% and 22.4%.

MICS 2025 estimates differed from DHS 2022 by 2.0 percentage points or less for all three indicators (p=0.50 to 0.91; S1 Table). MICS 2019 estimates were lower than DHS 2017-18, by 8.0 points for combined treatment (95% CI 1.6 to 14.4; p=0.014; S1 Fig). The DHS change in combined coverage between 2017-18 and 2022 was 0.3 points (−8.1 to 8.6). The difference between the MICS and DHS changes was uncertain (7.6 points, −1.6 to 16.8; p=0.11; S1 Table), over about six years in MICS and under five in the DHS. DHS ORS coverage fell from 83.3% to 73.8% (change −9.6 points, −16.3 to −2.9; S1 Table).

### Wealth and other characteristics

Combined treatment showed an unadjusted wealth gradient in both surveys (Table 2, Fig 1). Its SII was 10.6 points (1.5 to 19.7) in 2019 and 13.0 points (3.4 to 22.6) in 2025. The SII changed by 2.4 points (−10.8 to 15.7), and there was no evidence of non-linearity (p=0.81 in 2019 and 0.29 in 2025; S5 Table). The zinc and ORS gradients did not differ clearly in either survey (Table 2, S5 Table).

**Fig 1.**
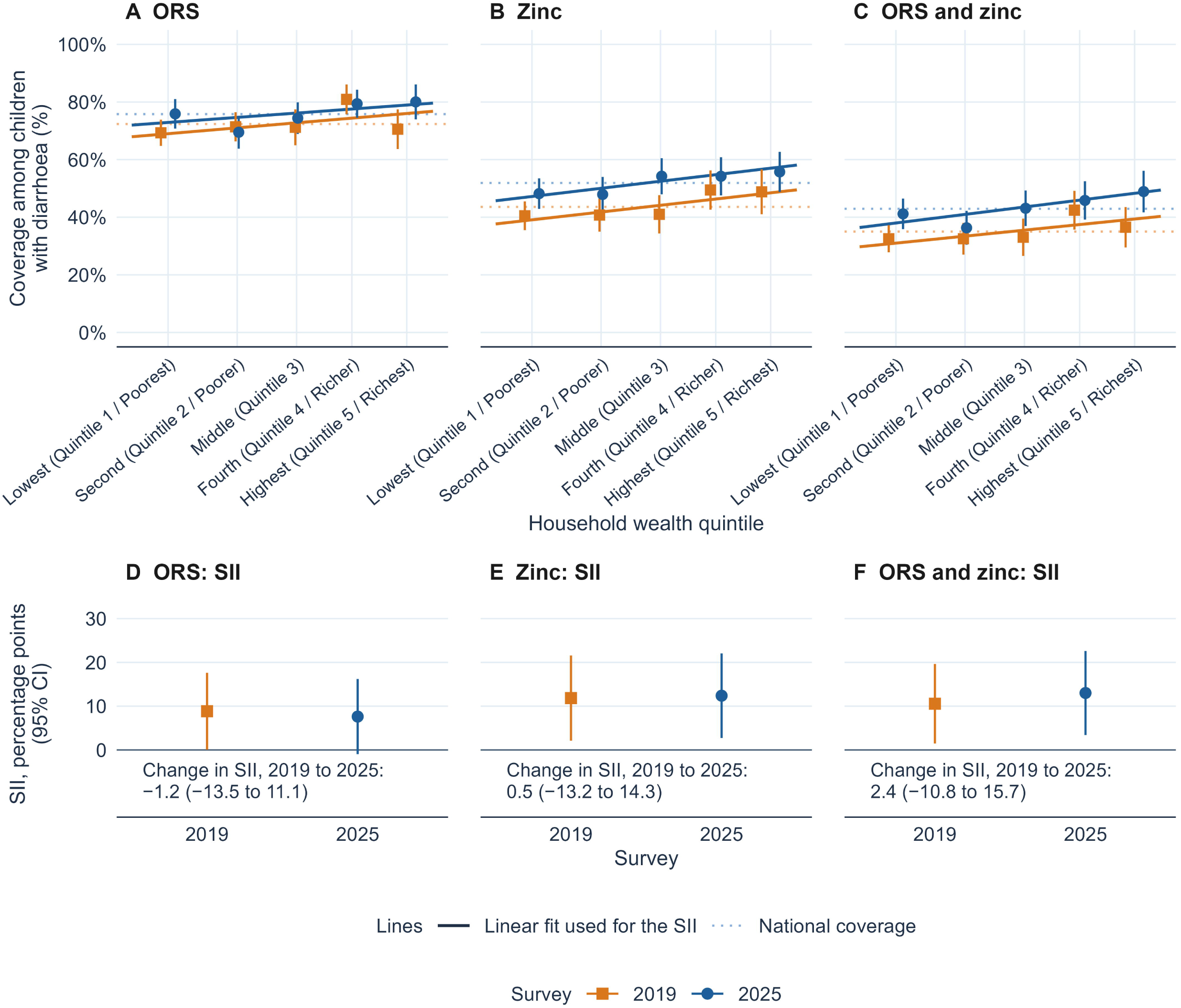
Coverage of ORS, zinc and combined treatment by household wealth quintile, MICS 2019 and 2025. The data are for children under five who had diarrhoea in the two weeks before each survey. In panels A to C, points show survey-weighted coverage in each wealth quintile in 2019 (orange squares) and 2025 (blue circles), with 95% confidence intervals. Each point sits at the quintile’s mean wealth rank among children with diarrhoea, shifted slightly so that points do not overlap. Solid lines show the linear fit that defines the slope index of inequality (SII), and dotted lines show national coverage. The SII is the absolute difference in coverage between the richest and poorest ends of the wealth distribution, in percentage points. Panels D to F show the SII with 95% confidence intervals and its change between surveys. Changes were computed from unrounded estimates, so they can differ by 0.1 from the difference between the rounded values. Overlapping intervals do not show that two estimates are similar.

Coverage did not rise steadily across quintiles in 2025, with 41.2% in the poorest, 36.3% in the second and 48.9% in the richest (Table 2, Fig 1). Model 1 showed no clear difference between the richest and poorest quintiles (aPR 1.03, 95% CI 0.86 to 1.24; Table 3). Adjusting for mother’s education, division, area, age and sex attenuated the pooled SII from 11.8 points (5.1 to 18.5) to 6.5 points (−1.7 to 14.6). This was a reduction of 5.3 points (0.3 to 10.3; S3 Table).

**Table 3.**
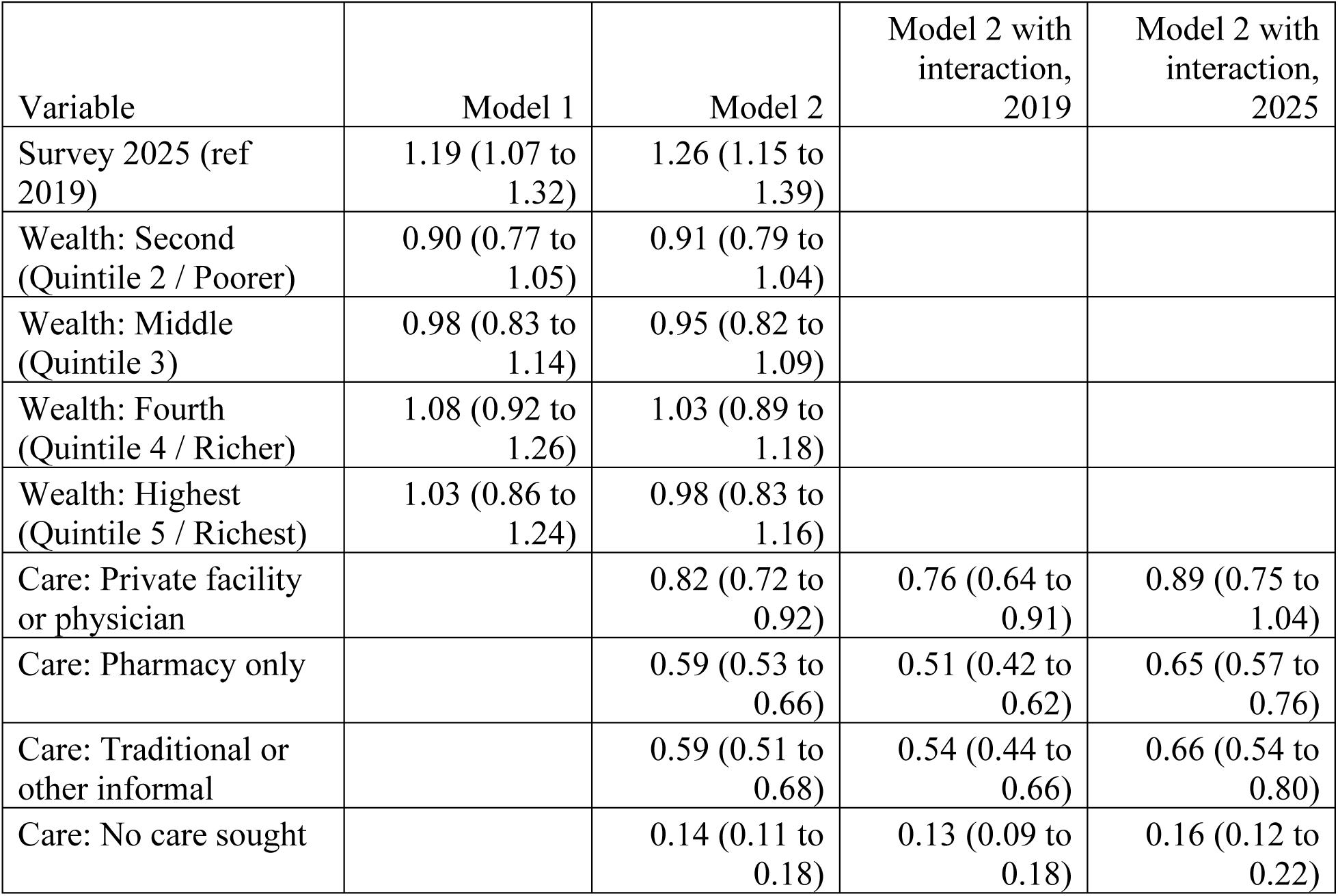
Adjusted prevalence ratios for combined ORS and zinc treatment by survey, wealth and reported source of care, pooled MICS 2019 and 2025 (n=3,222). Values are adjusted prevalence ratios (aPR) with 95% confidence intervals (CIs). The reference groups are the lowest wealth quintile (Quintile 1 / Poorest) and public health care. Pharmacy only means a private pharmacy and no other medical source, and no care sought means that no advice or treatment was sought.

| Variable | Model 1 | Model 2 | Model 2 with interaction, 2019 | Model 2 with interaction, 2025 |
|---|---|---|---|---|
| Survey 2025 (ref 2019) | 1.19 (1.07 to 1.32) | 1.26 (1.15 to 1.39) |  |  |
| Wealth: Second (Quintile 2 / Poorer) | 0.90 (0.77 to 1.05) | 0.91 (0.79 to 1.04) |  |  |
| Wealth: Middle (Quintile 3) | 0.98 (0.83 to 1.14) | 0.95 (0.82 to 1.09) |  |  |
| Wealth: Fourth (Quintile 4 / Richer) | 1.08 (0.92 to 1.26) | 1.03 (0.89 to 1.18) |  |  |
| Wealth: Highest (Quintile 5 / Richest) | 1.03 (0.86 to 1.24) | 0.98 (0.83 to 1.16) |  |  |
| Care: Private facility or physician |  | 0.82 (0.72 to 0.92) | 0.76 (0.64 to 0.91) | 0.89 (0.75 to 1.04) |
| Care: Pharmacy only |  | 0.59 (0.53 to 0.66) | 0.51 (0.42 to 0.62) | 0.65 (0.57 to 0.76) |
| Care: Traditional or other informal |  | 0.59 (0.51 to 0.68) | 0.54 (0.44 to 0.66) | 0.66 (0.54 to 0.80) |
| Care: No care sought |  | 0.14 (0.11 to 0.18) | 0.13 (0.09 to 0.18) | 0.16 (0.12 to 0.22) |

Model 1 gives the adjusted association of each characteristic with treatment. It includes survey year, wealth quintile, mother’s education (reference secondary), area (urban), division (Dhaka), age group (12 to 23 months) and sex (boys). Model 2 adds source of care, to compare sources. The last two columns add a survey-by-source interaction to Model 2 (F(4, 1988)=1.15, p=0.33). The 1,988 denominator degrees of freedom are the 2,017 design degrees of freedom minus the 29 model terms besides the intercept. All are survey-weighted Poisson models with a log link and linearised variance. S5 Table gives every coefficient, and Model 3 with illness proxies. In 2025, combined coverage ranged from 35.3% (95% CI 27.6 to 43.0) in Rangpur to 47.6% (40.5 to 54.6) in Dhaka division (Fig 2). The adjusted pooled test of differences between divisions gave p=0.050 (S5 Table). Sylhet and Barishal had the lowest adjusted ratios against Dhaka (aPR 0.75, 0.60 to 0.94, and 0.77, 0.63 to 0.93; S5 Table). Of 18 exploratory interaction tests, only zinc by sex had p<0.05 (p=0.004), and none passed a Bonferroni threshold (S5 Table).

**Fig 2.**
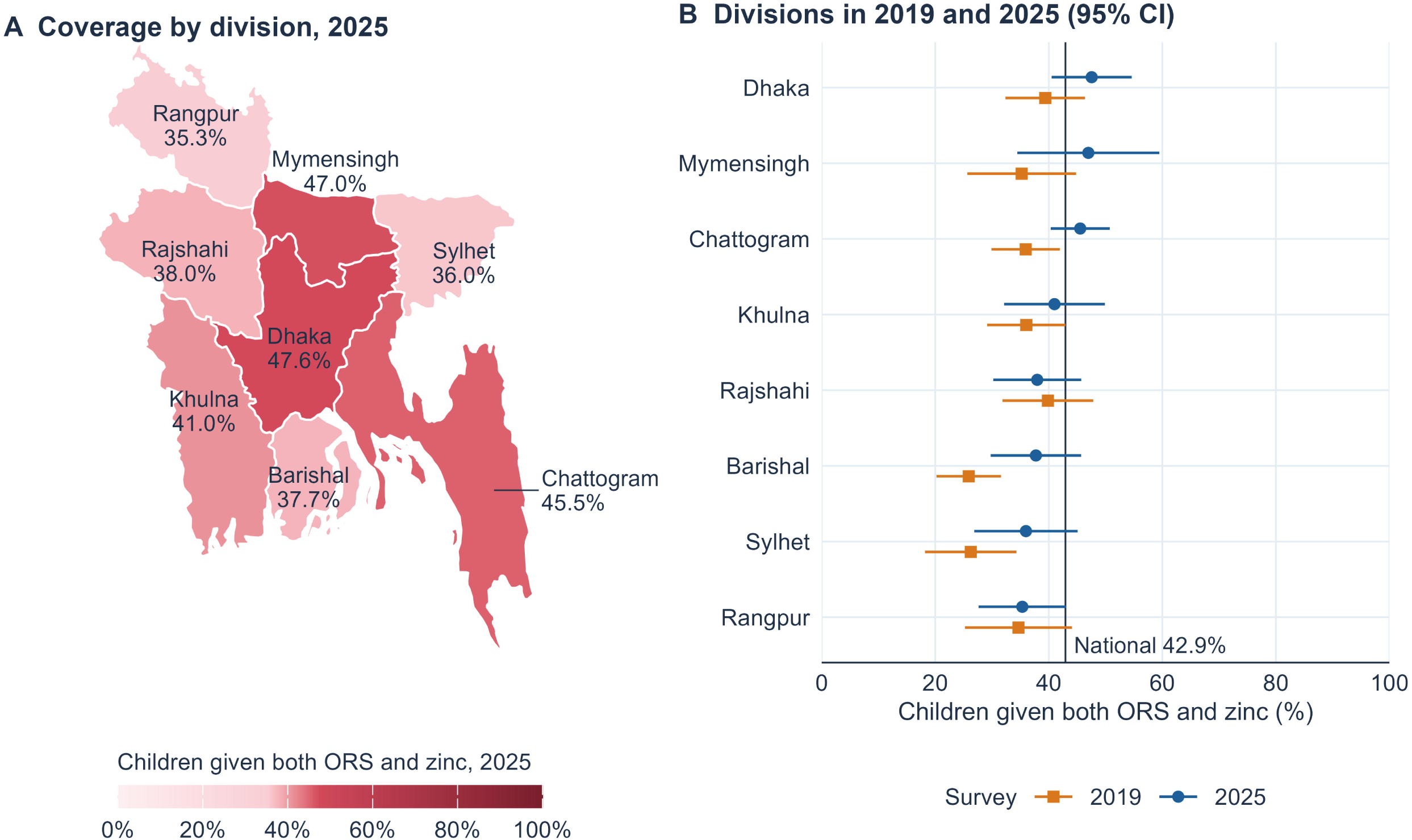
Combined ORS and zinc coverage by division, 2019 and 2025. Panel A shows the survey-weighted share of children with diarrhoea who received both ORS and zinc in each division in 2025. The colour scale runs from 0% to 100%, with its colour stops stretched over the observed range so that differences are visible. The stretch aids reading and does not mean that divisions differed. The map shows point estimates only, and Panel B gives their uncertainty. Panel B shows division estimates for 2019 (orange squares) and 2025 (blue circles) with 95% confidence intervals. The solid line marks the national estimate for 2025. Division boundaries are from geoBoundaries [43].

### Source of care

Any care was sought for 72.5% of children in 2019 and 71.5% in 2025 (change −1.1 points, 95% CI −4.7 to 2.5), but the reported places differed (Fig 3A). Pharmacy-only care rose from 25.9% (23.2 to 28.5) to 35.0% (32.4 to 37.6), a rise of 9.2 points (5.5 to 12.9; Table 1). Public, private facility and traditional or other informal care each fell slightly (Table 1). No care was sought for 27.5% of children in 2019 and 28.5% in 2025 (Table 1). Pharmacy-only care rose most in the middle quintile, by 16.1 points (7.6 to 24.5), and the poorest quintile, by 15.0 points (8.0 to 22.0; Fig 3A). The survey-by-quintile test gave p=0.079 (S5 Table).

**Fig 3.**
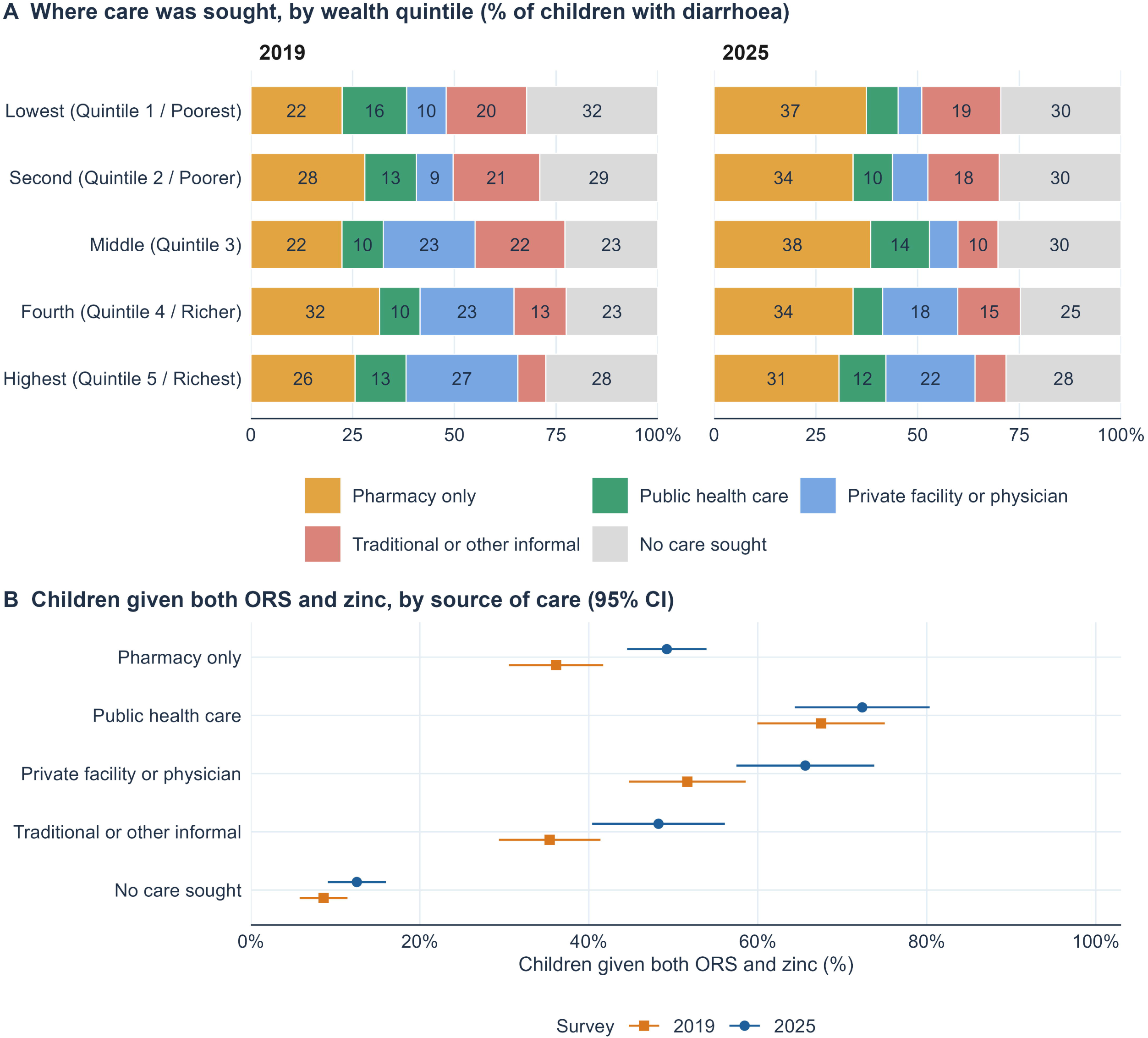
Reported source of care by wealth quintile and combined treatment coverage by source, 2019 and 2025. Panel A shows the survey-weighted share of children with diarrhoea in each source of care, by wealth quintile, in 2019 and 2025. Pharmacy-only care is shown first in each bar, and numbers give shares of 9% or more. Panel B shows the survey-weighted share of children who received both ORS and zinc in each source of care. Points show 2019 (orange squares) and 2025 (blue circles), with 95% confidence intervals. Children who named several places were assigned in the order public health care, private facility or physician, pharmacy only, traditional or other informal, and no care sought.

In 2025, combined coverage was 72.4% with public care, 65.6% with private facility or physician care and 49.2% with pharmacy-only care (Fig 3B). It was 48.2% with traditional or other informal care and 12.5% with no care reported. Against public care, pooled aPRs were 0.59 (95% CI 0.53 to 0.66) for pharmacy-only care and 0.14 (0.11 to 0.18) with no care reported (Table 3). The pharmacy-only aPR was 0.51 (0.42 to 0.62) in 2019 and 0.65 (0.57 to 0.76) in 2025, and evidence that these contrasts changed between surveys was weak (interaction p=0.33). In 2025, children with no care reported were 43.7% (95% CI 40.0 to 47.5) of children without combined treatment but 28.5% of all children (S4 Table). Pharmacy-only children were 31.2% (27.8 to 34.5) of children without combined treatment and 35.0% of all children (S4 Table). In 2019, pharmacies supplied 72.1% of the zinc and 63.9% of the ORS among children given them (S4 Table). Among children with no care reported, 37.7% (32.5 to 42.9) had ORS or zinc from a pharmacy (S4 Table).

Care from a public or private facility or physician rose with wealth (S4 Table). Its SII was 20.3 points (95% CI 11.2 to 29.5) in 2019 and 23.2 points (15.1 to 31.2) in 2025 (S4 Table). In the poorest quintile, this care fell from 25.6% to 13.6% (S4 Table). Yet combined coverage in the poorest quintile rose by 8.6 points (1.5 to 15.7), against 12.4 points (2.3 to 22.4) in the richest (Table 2). These gains did not differ clearly (difference 3.8 points, −8.3 to 15.9; S5 Table). Adding source of care attenuated the pooled SII by 5.6 points (2.6 to 8.6), or by 4.1 points (0.7 to 7.5) with other characteristics in the model (S3 Table). In the decomposition, change within sources added 9.7 points (6.0 to 13.4), and shifts across sources contributed −1.8 points (−3.4 to −0.2; S3 Table).

### Sensitivity analyses

Combined coverage rose in all five sensitivity analyses of the change, by 7.3 to 8.7 points (S5 Table). With quintile midpoints, the combined-treatment SII was 8.9 points (−0.4 to 18.3) in 2019 and 12.1 (2.2 to 22.0) in 2025, so the 2019 gradient is less certain. Ranking all children under five gave 10.5 and 13.2 points. Adding illness proxies changed the source aPRs by 0.01 or less, and other assignment orders and logistic regression with marginal standardisation gave pharmacy-only prevalence ratios of 0.60 to 0.61. With infants split, the aPR was 0.46 (95% CI 0.34 to 0.61) at 0 to 5 months and 1.07 (0.94 to 1.23) at 6 to 11 months. Both used 12 to 23 months as reference (S5 Table). The survey-year and source aPRs changed by 0.01 or less. Combined, the private physician, pharmacy-only and traditional groups were 59.9% of children in 2019 and 61.5% in 2025 (change 1.5 points, −2.4 to 5.4; S5 Table). Within them, the pharmacy-only share rose from 43.1% to 57.0%.

## Discussion

Combined ORS and zinc coverage among children with diarrhoea in Bangladesh rose between 2019 and 2025, but only 42.9% received both in 2025. Zinc was the larger gap, but one child in four received no ORS, and infants under six months had the lowest ORS coverage. An unadjusted wealth gradient in combined treatment was attenuated after adjustment for mother’s education, region and other characteristics. Coverage differed most by reported source of care. Reported pharmacy-only care became more common, and children with no care reported were over-represented among children without both treatments. Most children who missed both treatments were outside facility care, which shapes where programmes can reach them.

The 2025 level is about twice the global estimate of 22% [11]. A pooled estimate for twelve East African countries was 53% [37] and Nigeria reported 22% [38], although definitions differ. No division reached 50% in 2025, and even among children reported as public care, more than a quarter did not receive both. One third of children were given ORS without zinc, a gap that early SUZY monitoring also showed [15].

Infants under six months had the lowest ORS coverage. In Nigeria, young infants also received less zinc [38], but in Bangladesh they received zinc as often as older children, so their gap is mainly in ORS. Even so, about half of these young infants received no zinc. At this age, the Cochrane review found no clear benefit of zinc [5], and the 2004 WHO statement gave a lower dose, 10 mg rather than 20 mg daily [6]. Low reported ORS may partly reflect how caregivers recognise diarrhoea in young infants, and our data cannot judge clinical appropriateness. In a qualitative study in Dhaka, few mothers knew that ORS does not pass to the baby through breast milk [45]. Such beliefs may limit ORS use in breastfed infants, although this is untested.

Our results agree in part with a DHS study that found little difference in zinc coverage across wealth quintiles in 2022 [13]. For combined treatment, the adjusted contrast between extreme quintiles was also unclear in our data, but rank-based indices, which use every child [17,18], showed an unadjusted gradient. Adjustment for mother’s education, region and other characteristics attenuated the pooled gradient, so part of the gradient is shared with these characteristics. Wealth gaps in ORS and zinc have also been reported in India [46] and across 35 sub-Saharan African countries [47]. In Gujarat, India, gains in zinc use during scale-up were larger in the richest quintile [48]. Gains in our poorest and richest quintiles did not differ clearly. Reported pharmacy-only care became the largest single group by 2025 and rose most in poorer quintiles, in line with reports that poorer families use pharmacies and shops [22]. In 2019, pharmacies also supplied most zinc and ORS, including for children reported as facility care or no care. Where care is sought and where treatment is obtained are therefore different measures. Coverage was lower with pharmacy-only than with facility care. Because source of care was reported with treatment, this gap may reflect severity or the purchase itself, and it is not proof that pharmacies give poorer care.

Facility care fell by about half in the poorest quintile, yet the poorest still gained combined coverage. Coverage rose within private, pharmacy-only and traditional or other informal care, which may point to common factors such as supply, price, familiarity or demand. The rise may also partly reflect low MICS 2019 estimates, as discussed below. In sub-Saharan Africa, appropriate treatment was also more common after facility or community care than after informal care [21].

Children with no care reported were the largest group without combined treatment and rarely received zinc, although more than a third had ORS or zinc from a pharmacy in 2019. Community delivery through community clinics and NGO health workers is one option to test [12]. In a Ugandan trial, free ORS from community health workers raised ORS use by 19 percentage points over usual sales, with a larger effect for ORS with zinc [49]. Promoting zinc together with ORS was associated with more ORS use in a meta-analysis of low-to-moderate quality [50]. Reaching pharmacy-only children through drug shops is harder, because operators have limited training and weak regulation [26] and village doctors depend on medicine sales [24,27]. Programmes that only improve informal providers’ knowledge have had limited impact [28]. In seven Zambian public facilities, dispensing of both products rose after co-packaging in an uncontrolled comparison [51]. We found no test of co-packaging in South Asian drug shops. These findings fit proportionate universalism, in which action reaches everyone but is stronger where need is greater [52]. Action for all children could work through pharmacies, with extra effort for children with no care reported and young infants. These options need evaluation, which should also measure antibiotic use, out-of-pocket cost and referral of sick children.

The rise within MICS held in all five sensitivity analyses, but comparison with the DHS limits how firmly its size can be stated. MICS 2019 was below DHS 2017-18 for all three indicators, and this gap has several possible explanations, including sampling variability. MICS 2019 may be low, which would overstate the rise, or DHS 2017-18 may be high, which would not. Season, population covered and field practice differ between the programmes [44]. The DHS change in combined coverage was imprecise, and its interval included both a fall and a rise.

## Strengths and limitations

The surveys were nationally representative and asked the same treatment questions in 2019 and 2025, and we compared the estimates with the DHS using formal tests. A PubMed search on 1 November 2026 found no earlier national analysis in Bangladesh that compared ORS and zinc coverage across sources of care.

First, source of care was reported together with treatment, which is the main limitation. Buying ORS or zinc may lead a caregiver to report pharmacy care. Sicker children may be reported as facility care and treated more completely, and our illness proxies were crude. MICS 2025 did not ask where treatment was obtained, so this could not be checked. We therefore present the comparisons by source of care as descriptive.

Second, the place codes cannot separate village doctors from other groups, and the rise in pharmacy-only care may partly reflect how the codes were used. The item on the first place of care was asked of only 40 children in 2025, too few to check our order of assignment. Third, treatment was recalled over two weeks, some episodes were still under way, and caregivers may over-report recommended treatment. Fourth, coverage overstates correct treatment, because osmolarity, zinc dose and duration were not recorded. Fifth, wealth rank gives relative position within each survey, so one rank may not mean the same living standard in 2019 and 2025.

## Conclusions

Combined ORS and zinc coverage among children with diarrhoea in Bangladesh rose between 2019 and 2025, but only 42.9% received both. Zinc was the larger gap, one child in four received no ORS, and most infants under six months did not receive ORS. Unadjusted wealth inequality in combined coverage was attenuated after adjustment for mother’s education, region and other characteristics. Coverage also differed by reported source of care, being highest with public health care and lowest with no care reported. Children with no care reported were over-represented among those without both treatments. Programmes through pharmacies and for these children need evaluation. Surveys should again ask where ORS and zinc were obtained, so that coverage by source of care can be validated.

## Data Availability

The Multiple Indicator Cluster Survey data sets for Bangladesh (2006, 2012-13, 2019 and 2025) are available from UNICEF on registration at https://mics.unicef.org/surveys. The Demographic and Health Survey data sets for Bangladesh (2004 to 2022) are available from The DHS Program on registration at https://dhsprogram.com. Other researchers can obtain the data in the same way as the authors, and the authors had no special access privileges.

## Acknowledgments

We thank the Bangladesh Bureau of Statistics and UNICEF for access to the Multiple Indicator Cluster Survey data. We also thank The DHS Program for access to the Demographic and Health Survey data.

## Author contributions

SMAUH conceived the study and designed the methodology. He curated the data, performed the formal analysis, and created the visualizations. He also wrote the original draft, revised the manuscript, and managed the project. MKH contributed to data curation, wrote part of the original draft, and reviewed and edited the manuscript. MKI contributed to interpretation of the results and reviewed the manuscript. MAR, TA, MRI and SMRH contributed to interpretation of the results and reviewed the manuscript. KUB supervised the study, validated the results, and reviewed and edited the manuscript. All authors read and approved the final manuscript.

---

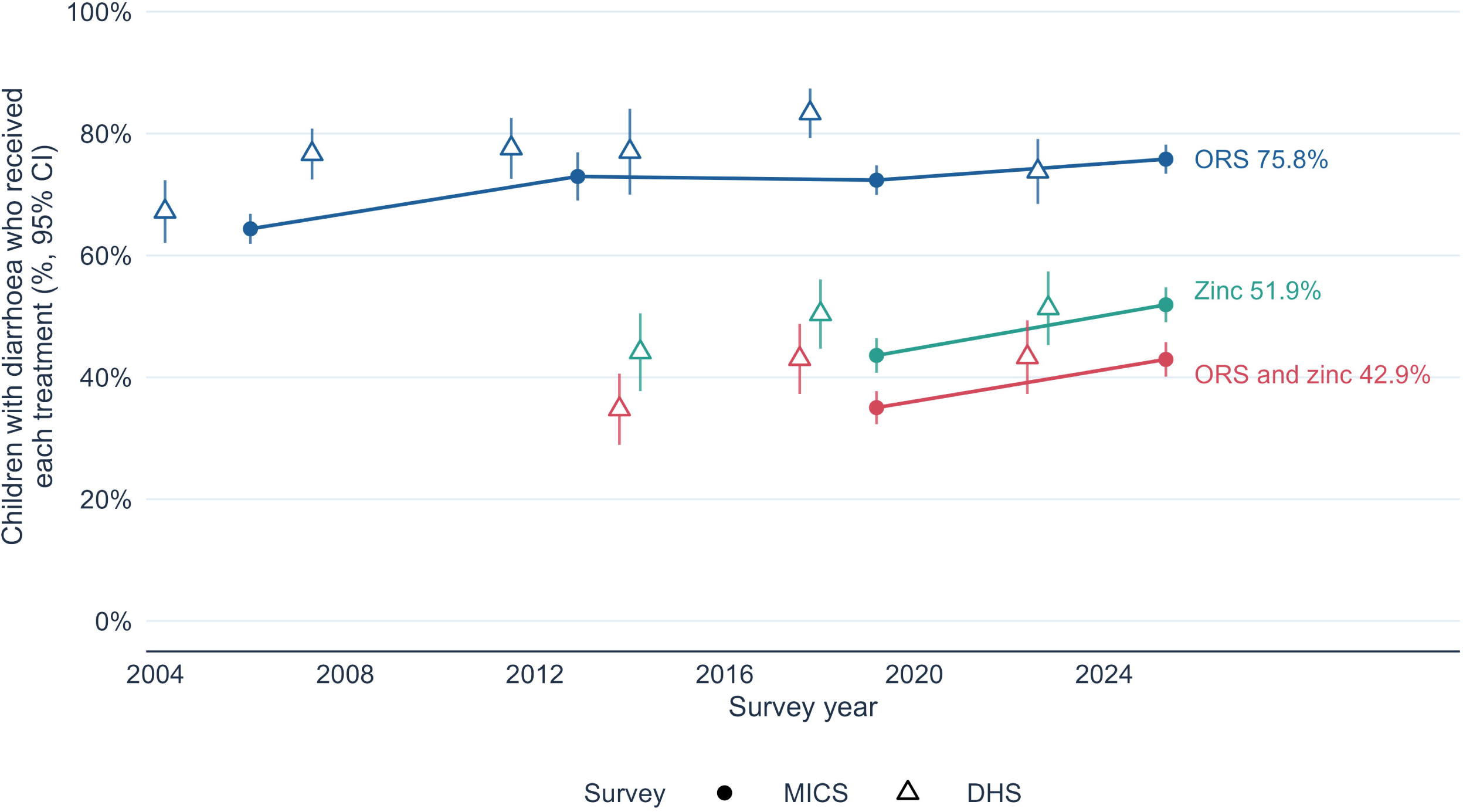

## Supporting information

**S1 Fig. National coverage of ORS, zinc and combined treatment in MICS and DHS, 2004 to 2025.** (TIF)

**S1 Table. Questions, definitions and places of care by survey, reproduction of published estimates, and comparison with Demographic and Health Survey estimates. (DOCX)**

**S2 Table. Coverage with confidence intervals and numbers of children by characteristic, and slope index of inequality for each treatment. (DOCX)**

**S3 Table. Wealth gradient under adjustment, and decomposition of the change in combined coverage. (DOCX)**

**S4 Table. Treatment combinations, care from facilities and physicians by wealth, and where ORS and zinc were obtained. (DOCX)**

**S5 Table. Sensitivity analyses, full models and further checks. (DOCX)**

**S1 Checklist. STROBE checklist for cross-sectional studies.** (DOCX)

