## Supplementary material for "Oral rehydration salts and zinc for childhood diarrhoea in Bangladesh, 2019 and 2025: coverage, wealth inequality and reported source of care": S1_Table.docx

### **S1 Table. Questions, definitions and places of care by survey, reproduction of published estimates, and comparison with Demographic and Health Survey estimates**

| Item | 2006 | 2012-13 | 2019 | 2025 |
| --- | --- | --- | --- | --- |
| Diarrhoea in past two weeks | CA1 | CA1 | CA1 | DA1 |
| Fluid from ORS packet | CA2A | CA4A | CA7A (packet saline) | DA9A |
| Second ORS item | CA2C (pre-packaged ORS fluid) | CA4B (rice-based ORS packet) | CA7B (rice saline) | DA9B (rice-based ORS) |
| Zinc | Not asked | CA6C, recorded only if the caregiver said something else was given (CA5) and named zinc | CA7C, direct question | DA9C, direct question |
| Care sought for diarrhoea | Not asked for diarrhoea | Not asked for diarrhoea | CA5, places CA6 | DA5, places DA6 |
| Where ORS and zinc were obtained | Not asked | Not asked | CA9, CA11 | Not asked |
| Severity proxies (sensitivity analysis) |  |  | CA3, CA4, CA16 | DA3, DA4, AR1 |

A. Questions, definitions and places of care by survey.

Any ORS was either ORS item. “Don’t know” answers counted as not given (six for zinc and two for ORS in 2019, and two for each in 2025). Less to drink or eat included “much less”, “somewhat less” and “nothing” or “stopped food”. Source of care had five groups, assigned in this order: public; private facility or physician; pharmacy only; traditional or other informal; no care sought.

Places where care was sought, by survey (weighted percent of children with diarrhoea who named each place, unweighted number in brackets). Several places could be named. “Not listed” means the code did not exist in that survey. There is no code for village doctors in either survey.

| Care group | Place | 2019 | 2025 |
| --- | --- | --- | --- |
| Public health care | Government hospital | 10.0 (157) | 8.0 (135) |
| Public health care | Government health centre | 1.4 (20) | 1.2 (23) |
| Public health care | Community clinic | 1.2 (21) | 0.3 (5) |
| Public health care | Government community health worker | 0.3 (7) | 0.4 (8) |
| Public health care | Mobile or outreach clinic | 0.0 (0) | 0.0 (0) |
| Public health care | Other public | 0.0 (0) | 0.2 (2) |
| Private facility or physician | Private hospital or clinic | 3.1 (49) | 3.1 (51) |
| Private facility or physician | Private physician | 12.0 (197) | 8.7 (143) |
| Private facility or physician | NGO community health worker | 1.5 (20) | 0.0 (0) |
| Private facility or physician | Private mobile clinic | 0.0 (1) | 0.2 (4) |
| Private facility or physician | NGO clinic or hospital | 0.7 (9) | not listed |
| Private facility or physician | Other private medical | 0.1 (3) | 0.0 (0) |
| Private facility or physician | DK public or private | 0.0 (0) | not listed |
| Private facility or physician | NGO hospital | not listed | 0.0 (0) |
| Private facility or physician | NGO clinic | not listed | 0.3 (7) |
| Private facility or physician | Other NGO | not listed | 0.0 (0) |
| Private facility or physician | DK public, private or NGO | not listed | 0.1 (2) |
| Pharmacy | Private pharmacy | 27.7 (397) | 35.8 (606) |
| Traditional | Traditional practitioner | 15.9 (261) | 11.7 (176) |
| Other informal | Relative or friend | 0.2 (4) | 0.4 (7) |
| Other informal | Shop, market or street | 2.0 (46) | 2.3 (35) |
| Other informal | Other | 1.2 (21) | 0.9 (16) |
| Other informal | DK | not listed | 0.1 (1) |

### B. Reproduction of published survey estimates, reconciliation of the combined indicator, and comparison with Demographic and Health Survey estimates.

Percent of children under five with diarrhoea in the past two weeks. Report values are from the MICS survey findings reports. The 2012-13 zinc item was recorded only after an unprompted “anything else given” question and is not comparable with later surveys. The manuscript uses any ORS in both 2019 and 2025.

| Survey | Indicator | Report | This analysis (95% CI) |
| --- | --- | --- | --- |
| 2006 | Fluid from ORS packet | 62.7 | 62.7 (60.2 to 65.2) |
| 2006 | Pre-packaged or rice-based ORS | 3.7 | 3.7 (2.6 to 4.9) |
| 2012 | Any ORS | 73.0 | 73.0 (69.0 to 76.9) |
| 2012 | Zinc | 11.8 | 12.1 (8.7 to 15.5) |
| 2019 | Fluid from ORS packet | 71.1 | 71.1 (68.6 to 73.6) |
| 2019 | Pre-packaged or rice-based ORS | 10.9 | 10.9 (9.1 to 12.6) |
| 2019 | Any ORS | 72.4 | 72.4 (69.9 to 74.8) |
| 2019 | Zinc | 43.6 | 43.6 (40.7 to 46.4) |
| 2019 | ORS and zinc | 35.0 | 35.0 (32.3 to 37.7) |
| 2025 | Fluid from ORS packet | 74.2 | 74.2 (71.8 to 76.6) |
| 2025 | Pre-packaged or rice-based ORS | 15.4 | 15.4 (13.4 to 17.5) |
| 2025 | Any ORS | 75.8 | 75.8 (73.4 to 78.2) |
| 2025 | Zinc | 51.9 | 51.9 (49.0 to 54.8) |
| 2025 | ORS and zinc | 41.6 | 42.9 (40.1 to 45.8) |

Reconciliation of the combined indicator (weighted percent of children with diarrhoea, 95% CI). Report indicator TC.13a counts any ORS. The report defines TC.13b as ORS and zinc, with no packet rule. The 2019 report value (35.0%) matches any ORS and zinc. The 2025 report value (41.6%) was reproduced only with packet ORS, in total and in every sex, area, wealth and division row checked. We cannot see the calculation in the report. The 1.3-point gap in 2025 equals the children given rice-based ORS alone with zinc. The change between surveys is 7.9 points with any ORS, 7.3 with packet ORS, and 6.6 if 35.0% is set beside 41.6%. The rice-based item is the column the reports label pre-packaged.

| Measure | 2019 | 2025 |
| --- | --- | --- |
| Packet ORS | 71.1 (68.6 to 73.6) | 74.2 (71.8 to 76.6) |
| Rice-based ORS | 10.9 (9.1 to 12.6) | 15.4 (13.4 to 17.5) |
| Both ORS types | 9.6 (7.9 to 11.3) | 13.8 (11.9 to 15.8) |
| Packet ORS only | 61.5 (58.8 to 64.2) | 60.4 (57.6 to 63.1) |
| Rice-based ORS only | 1.2 (0.7 to 1.8) | 1.6 (0.8 to 2.4) |
| Any ORS (TC.13a) | 72.4 (69.9 to 74.8) | 75.8 (73.4 to 78.2) |
| Zinc | 43.6 (40.7 to 46.4) | 51.9 (49.0 to 54.8) |
| Packet ORS and zinc | 34.3 (31.6 to 37.0) | 41.6 (38.8 to 44.5) |
| Rice-based ORS and zinc | 7.1 (5.7 to 8.6) | 10.0 (8.4 to 11.7) |
| Both ORS types and zinc | 6.5 (5.1 to 7.8) | 8.7 (7.2 to 10.3) |
| Rice-based ORS only and zinc | 0.7 (0.2 to 1.2) | 1.3 (0.6 to 2.0) |
| Any ORS and zinc (this study) | 35.0 (32.3 to 37.7) | 42.9 (40.1 to 45.8) |

DHS estimates used as a validity check. The DHS Program publishes point estimates only for these indicators. We re-estimated them from the DHS children’s recode files with survey weights, clusters and strata. The denominator was living children born in the five years before the survey with diarrhoea in the past two weeks. ORS meant fluid from an ORS packet or pre-packaged ORS, and zinc meant zinc tablets or syrup.

| Survey | Indicator | Children with diarrhoea | Published | This analysis (95% CI) |
| --- | --- | --- | --- | --- |
| DHS 2004 | ORS | 485 | 67.1 | 67.2 (62.1 to 72.3) |
| DHS 2007 | ORS | 560 | 76.6 | 76.6 (72.5 to 80.8) |
| DHS 2011 | ORS | 395 | 77.6 | 77.6 (72.6 to 82.6) |
| DHS 2014 | ORS | 371 | 77.0 | 77.0 (70.0 to 84.1) |
| DHS 2014 | Zinc | 371 | 44.1 | 44.1 (37.7 to 50.5) |
| DHS 2014 | ORS and zinc | 371 | 34.8 | 34.8 (28.9 to 40.6) |
| DHS 2017-18 | ORS | 412 | 83.3 | 83.3 (79.3 to 87.4) |
| DHS 2017-18 | Zinc | 412 | 50.4 | 50.4 (44.7 to 56.1) |
| DHS 2017-18 | ORS and zinc | 412 | 43.0 | 43.0 (37.3 to 48.8) |
| DHS 2022 | ORS | 410 | 73.8 | 73.8 (68.4 to 79.1) |
| DHS 2022 | Zinc | 410 | 51.3 | 51.3 (45.3 to 57.4) |
| DHS 2022 | ORS and zinc | 410 | 43.3 | 43.3 (37.3 to 49.3) |

Comparison of MICS and DHS estimates. Differences are in percentage points, from z tests that treat the surveys as independent samples. Standard errors were derived from the 95% confidence intervals. With these sample sizes, the DHS could detect a change in combined treatment of about 12 percentage points or more between 2017-18 and 2022 (80% power, 5% significance). MICS interviews took place from January to June 2019 and from February to June 2025. DHS interviews took place from October 2017 to March 2018 and mostly from August to December 2022.

| Comparison | Indicator | Estimates (%) | Difference (95% CI) | p |
| --- | --- | --- | --- | --- |
| MICS 2025 vs DHS 2022 | ORS | 75.8 vs 73.8 | 2.0 (-3.8 to 7.9) | 0.50 |
| MICS 2025 vs DHS 2022 | Zinc | 51.9 vs 51.3 | 0.6 (-6.1 to 7.3) | 0.87 |
| MICS 2025 vs DHS 2022 | ORS and zinc | 42.9 vs 43.3 | -0.4 (-7.0 to 6.3) | 0.91 |
| MICS 2019 vs DHS 2017-18 | ORS | 72.4 vs 83.3 | -11.0 (-15.7 to -6.3) | <0.001 |
| MICS 2019 vs DHS 2017-18 | Zinc | 43.6 vs 50.4 | -6.8 (-13.1 to -0.4) | 0.037 |
| MICS 2019 vs DHS 2017-18 | ORS and zinc | 35.0 vs 43.0 | -8.0 (-14.4 to -1.6) | 0.014 |
| DHS 2022 vs DHS 2017-18 | ORS | 73.8 vs 83.3 | -9.6 (-16.3 to -2.9) | 0.005 |
| DHS 2022 vs DHS 2017-18 | Zinc | 51.3 vs 50.4 | 1.0 (-7.3 to 9.3) | 0.82 |
| DHS 2022 vs DHS 2017-18 | ORS and zinc | 43.3 vs 43.0 | 0.3 (-8.1 to 8.6) | 0.95 |

Change versus change: the MICS rise in combined coverage from 2019 to 2025 was 7.9 points (4.0 to 11.8). The DHS change from 2017-18 to 2022 was 0.3 points (-8.1 to 8.6). The difference between the two changes was 7.6 points (-1.6 to 16.8; p=0.11). The MICS 2025 minus DHS 2022 difference for combined treatment was -0.4 points (-7.0 to 6.3). The data are compatible with differences of up to about 7 points in either direction, and they do not show agreement within a tighter margin.

Packet ORS only: the DHS ORS item is fluid from a packet, and adding pre-packaged ORS changed no DHS value. With packet ORS only, MICS combined coverage was 34.3% (31.6 to 37.0) in 2019 and 41.6% (38.8 to 44.5) in 2025. The DHS values were 43.0% (37.2 to 48.8) in 2017-18 and 43.3% (37.2 to 49.4) in 2022.

Prevalence and season. Diarrhoea prevalence among living children under five was 4.7% (4.2 to 5.2) in DHS 2017-18 and 4.8% (4.2 to 5.4) in DHS 2022. In MICS 2019 and 2025 it was 6.9% (6.5 to 7.3) and 7.1% (6.7 to 7.5). DHS cases came from January to March and October to December in 2017-18. In 2022 they came mostly from August to December. Only 103 DHS 2017-18 cases and no 2022 cases fell in February to May, the months sampled in both MICS rounds, so a season-matched DHS comparison was not possible.
