## Supplementary material for "Oral rehydration salts and zinc for childhood diarrhoea in Bangladesh, 2019 and 2025: coverage, wealth inequality and reported source of care": S2_Table.docx

### **S2 Table. Coverage with confidence intervals and numbers of children by characteristic, and slope index of inequality for each treatment**

Coverage among children under five with diarrhoea (weighted percent, 95% CI), with unweighted numbers of children. Small groups, such as Mymensingh, have wide intervals.

| Characteristic | Group | n 2019 | n 2025 | ORS 2019 | ORS 2025 | Zinc 2019 | Zinc 2025 | ORS and zinc 2019 | ORS and zinc 2025 |
| --- | --- | --- | --- | --- | --- | --- | --- | --- | --- |
| Total | Total | 1,589 | 1,634 | 72.4 (69.9 to 74.8) | 75.8 (73.4 to 78.2) | 43.6 (40.7 to 46.4) | 51.9 (49.0 to 54.8) | 35.0 (32.3 to 37.7) | 42.9 (40.1 to 45.8) |
| Sex | Male | 852 | 873 | 72.8 (69.5 to 76.1) | 76.0 (72.7 to 79.2) | 45.3 (41.4 to 49.2) | 47.4 (43.5 to 51.3) | 35.8 (32.1 to 39.5) | 40.6 (36.7 to 44.4) |
|  | Female | 737 | 761 | 71.9 (68.3 to 75.4) | 75.6 (72.2 to 79.0) | 41.6 (37.5 to 45.7) | 56.9 (52.9 to 61.0) | 34.1 (30.1 to 38.2) | 45.6 (41.6 to 49.6) |
| Age (months) | 0-5 | 155 | 134 | 28.0 (20.6 to 35.5) | 37.0 (27.6 to 46.3) | 43.0 (34.3 to 51.8) | 50.1 (40.3 to 59.8) | 15.4 (9.2 to 21.5) | 22.4 (14.0 to 30.7) |
|  | 6-11 | 254 | 285 | 70.5 (64.2 to 76.7) | 76.1 (70.1 to 82.1) | 49.9 (42.8 to 57.0) | 60.8 (54.3 to 67.2) | 38.7 (31.9 to 45.6) | 50.0 (43.1 to 56.8) |
|  | 12-23 | 459 | 483 | 76.0 (71.7 to 80.3) | 75.4 (71.0 to 79.7) | 46.2 (41.0 to 51.3) | 51.9 (46.6 to 57.2) | 38.2 (33.1 to 43.2) | 44.8 (39.6 to 50.0) |
|  | 24-35 | 330 | 337 | 79.0 (73.7 to 84.2) | 81.1 (76.1 to 86.0) | 43.8 (37.7 to 49.9) | 50.6 (44.5 to 56.6) | 39.4 (33.3 to 45.4) | 44.0 (37.9 to 50.1) |
|  | 36-47 | 232 | 222 | 82.0 (76.2 to 87.7) | 84.1 (78.5 to 89.7) | 43.0 (36.2 to 49.9) | 48.5 (40.7 to 56.3) | 39.0 (32.3 to 45.8) | 43.1 (35.4 to 50.9) |
|  | 48-59 | 158 | 173 | 81.7 (74.8 to 88.7) | 87.2 (81.8 to 92.5) | 26.5 (18.0 to 35.1) | 45.2 (36.7 to 53.7) | 24.3 (15.9 to 32.8) | 40.1 (31.7 to 48.6) |
| Area | Urban | 299 | 409 | 70.9 (65.3 to 76.6) | 79.6 (75.3 to 83.8) | 44.3 (37.6 to 50.9) | 56.3 (50.3 to 62.3) | 33.8 (27.5 to 40.1) | 48.1 (42.2 to 53.9) |
|  | Rural | 1,290 | 1,225 | 72.7 (70.1 to 75.4) | 74.4 (71.5 to 77.3) | 43.4 (40.3 to 46.5) | 50.3 (47.0 to 53.5) | 35.3 (32.4 to 38.3) | 41.0 (37.8 to 44.2) |
| Division | Barishal | 258 | 171 | 71.9 (65.9 to 78.0) | 70.5 (62.3 to 78.6) | 32.3 (26.5 to 38.1) | 47.1 (39.3 to 54.9) | 25.9 (20.2 to 31.6) | 37.7 (29.8 to 45.7) |
|  | Chattogram | 385 | 491 | 69.2 (63.9 to 74.4) | 78.0 (73.6 to 82.5) | 44.8 (38.5 to 51.1) | 53.9 (48.7 to 59.1) | 35.9 (29.9 to 42.0) | 45.5 (40.3 to 50.7) |
|  | Dhaka | 249 | 290 | 79.6 (74.0 to 85.3) | 77.5 (72.3 to 82.7) | 48.6 (41.2 to 55.9) | 57.2 (50.1 to 64.4) | 39.4 (32.4 to 46.4) | 47.6 (40.5 to 54.6) |
|  | Khulna | 204 | 168 | 78.2 (72.3 to 84.1) | 76.3 (69.9 to 82.7) | 43.2 (35.7 to 50.6) | 52.4 (43.1 to 61.8) | 36.0 (29.1 to 42.9) | 41.0 (32.1 to 49.9) |
|  | Mymensingh | 113 | 87 | 66.5 (57.8 to 75.2) | 82.0 (72.8 to 91.3) | 40.5 (30.6 to 50.4) | 51.2 (39.3 to 63.2) | 35.2 (25.6 to 44.8) | 47.0 (34.5 to 59.5) |
|  | Rajshahi | 152 | 155 | 75.9 (68.9 to 82.9) | 73.1 (66.0 to 80.2) | 48.4 (40.4 to 56.3) | 44.8 (36.5 to 53.1) | 39.8 (31.8 to 47.8) | 38.0 (30.2 to 45.7) |
|  | Rangpur | 116 | 164 | 68.7 (59.6 to 77.9) | 71.5 (63.4 to 79.5) | 51.2 (41.4 to 61.0) | 48.0 (39.4 to 56.5) | 34.7 (25.2 to 44.1) | 35.3 (27.6 to 43.0) |
|  | Sylhet | 112 | 108 | 62.1 (53.1 to 71.1) | 66.6 (56.1 to 77.2) | 34.4 (25.2 to 43.5) | 46.4 (37.5 to 55.3) | 26.3 (18.2 to 34.3) | 36.0 (26.9 to 45.1) |
| Mother’s education | No education | 201 | 93 | 69.5 (62.0 to 77.0) | 78.7 (69.5 to 87.9) | 39.9 (31.6 to 48.2) | 49.4 (37.8 to 61.0) | 31.4 (23.4 to 39.4) | 43.0 (31.4 to 54.5) |
|  | Primary | 425 | 332 | 72.5 (67.8 to 77.2) | 73.5 (68.2 to 78.8) | 39.2 (33.9 to 44.5) | 47.4 (41.3 to 53.4) | 30.9 (25.8 to 36.0) | 36.9 (31.1 to 42.8) |
|  | Secondary | 758 | 879 | 71.6 (68.0 to 75.2) | 73.9 (70.4 to 77.4) | 46.2 (42.3 to 50.2) | 52.0 (48.0 to 55.9) | 37.5 (33.7 to 41.2) | 42.6 (38.7 to 46.5) |
|  | Higher secondary or more | 205 | 330 | 77.5 (71.4 to 83.7) | 82.5 (78.1 to 86.8) | 45.5 (38.1 to 53.0) | 57.0 (50.7 to 63.3) | 37.2 (29.7 to 44.7) | 49.8 (43.3 to 56.2) |
| Wealth quintile | Lowest (Quintile 1 / Poorest) | 489 | 406 | 69.3 (64.8 to 73.9) | 75.9 (70.8 to 81.0) | 40.5 (35.5 to 45.5) | 48.2 (42.9 to 53.5) | 32.5 (27.8 to 37.3) | 41.2 (35.9 to 46.5) |
|  | Second (Quintile 2 / Poorer) | 379 | 342 | 71.4 (66.3 to 76.4) | 69.5 (63.8 to 75.2) | 40.8 (35.0 to 46.5) | 47.9 (41.9 to 54.0) | 32.5 (27.0 to 38.0) | 36.3 (30.5 to 42.2) |
|  | Middle (Quintile 3) | 257 | 332 | 71.2 (65.0 to 77.4) | 74.4 (68.9 to 79.9) | 41.0 (34.4 to 47.7) | 54.2 (48.0 to 60.5) | 33.0 (26.6 to 39.5) | 43.1 (36.9 to 49.3) |
|  | Fourth (Quintile 4 / Richer) | 267 | 297 | 80.9 (75.7 to 86.1) | 79.4 (74.5 to 84.3) | 49.5 (42.6 to 56.3) | 54.2 (47.6 to 60.8) | 42.4 (35.7 to 49.2) | 45.8 (39.2 to 52.5) |
|  | Highest (Quintile 5 / Richest) | 196 | 257 | 70.6 (63.7 to 77.4) | 80.0 (74.0 to 86.1) | 48.8 (41.0 to 56.6) | 55.7 (48.8 to 62.7) | 36.5 (29.5 to 43.5) | 48.9 (41.7 to 56.1) |
| Source of care | Public health care | 201 | 172 | 87.7 (82.6 to 92.8) | 91.9 (87.4 to 96.5) | 75.5 (68.5 to 82.5) | 77.8 (70.3 to 85.4) | 67.5 (59.9 to 75.0) | 72.4 (64.4 to 80.3) |
|  | Private facility or physician | 270 | 202 | 78.3 (72.5 to 84.1) | 80.8 (74.6 to 86.9) | 64.3 (57.6 to 71.0) | 79.8 (73.4 to 86.1) | 51.7 (44.8 to 58.6) | 65.6 (57.5 to 73.8) |
|  | Pharmacy only | 368 | 592 | 77.2 (72.5 to 82.0) | 76.5 (72.6 to 80.4) | 45.2 (39.3 to 51.2) | 62.3 (57.8 to 66.9) | 36.1 (30.5 to 41.7) | 49.2 (44.5 to 53.9) |
|  | Traditional or other informal | 303 | 213 | 72.5 (66.9 to 78.1) | 79.2 (73.6 to 84.7) | 47.0 (40.7 to 53.3) | 57.8 (50.0 to 65.6) | 35.4 (29.4 to 41.4) | 48.2 (40.4 to 56.1) |
|  | No care sought | 447 | 455 | 57.0 (51.9 to 62.1) | 65.5 (60.4 to 70.7) | 12.6 (9.2 to 16.0) | 15.2 (11.4 to 19.0) | 8.6 (5.8 to 11.4) | 12.5 (9.1 to 16.0) |

Slope index of inequality by treatment and survey (percentage points, 95% CI). It is the predicted difference in coverage between the richest and poorest ends of the wealth distribution.

| Index | ORS 2019 | ORS 2025 | Zinc 2019 | Zinc 2025 | ORS and zinc 2019 | ORS and zinc 2025 |
| --- | --- | --- | --- | --- | --- | --- |
| Slope index (points) | 8.8 (0.1 to 17.6) | 7.6 (-1.0 to 16.2) | 11.8 (2.1 to 21.6) | 12.4 (2.7 to 22.1) | 10.6 (1.5 to 19.7) | 13.0 (3.4 to 22.6) |
