## Supplementary material for "Oral rehydration salts and zinc for childhood diarrhoea in Bangladesh, 2019 and 2025: coverage, wealth inequality and reported source of care": S3_Table.docx

### **S3 Table. Wealth gradient under adjustment, and decomposition of the change in combined coverage**

Slope index of inequality for combined treatment in pooled data, with survey year, under different adjustment sets (percentage points, 95% CI from 1,000 subbootstrap replicate weights). Characteristics are mother’s education, division, area, child age and sex. Illness proxies are less to drink, less to eat and cough. Each reduction compares the models without and with the named set of covariates. These are statistical attenuations, not mediation estimates.

| Adjustment | Slope index or reduction, points (95% CI) |
| --- | --- |
| None | 11.8 (5.1 to 18.5) |
| Characteristics | 6.5 (-1.7 to 14.6) |
| Care group | 6.2 (-0.1 to 12.5) |
| Characteristics and care group | 2.4 (-5.4 to 10.2) |
| Characteristics, care group and illness proxies | 2.5 (-5.3 to 10.3) |
| Reduction from care group | 5.6 (2.6 to 8.6) |
| Reduction from care group after characteristics | 4.1 (0.7 to 7.5) |
| Reduction from characteristics | 5.3 (0.3 to 10.3) |

Decomposition of the 2019 to 2025 change in combined coverage (percentage points, 95% CI from 1,000 subbootstrap replicate weights). For sources k, with share S and coverage C in each survey, the shift part is the sum of (S2025k - S2019k) x (C2019k + C2025k) / 2. The within part is the sum of (C2025k - C2019k) x (S2019k + S2025k) / 2. The sources are exclusive and exhaustive, so the two parts add to the total. A negative shift part means that the share of children fell in sources with higher coverage. It does not show that individual children moved between sources. The decomposition is descriptive and not adjusted for case mix. The total change has a slightly different CI in the main text (4.0 to 11.8), which comes from Taylor series linearisation. With two other random seeds, the total ran from 4.0 to 11.9 and from 4.0 to 11.8, so the intervals were stable.

| Component | Points (95% CI) |
| --- | --- |
| Total change | 7.9 (3.9 to 11.9) |
| Change within sources of care (all) | 9.7 (6.0 to 13.4) |
| Within: public health care | 0.6 (-0.7 to 1.9) |
| Within: private facility or physician | 2.0 (0.4 to 3.6) |
| Within: pharmacy only | 4.0 (1.7 to 6.3) |
| Within: traditional or other informal | 2.0 (0.5 to 3.5) |
| Within: no care sought | 1.1 (-0.2 to 2.4) |
| Shift between sources of care (all) | -1.8 (-3.4 to -0.2) |
| Shift: public health care | -1.8 (-3.6 to -0.1) |
| Shift: private facility or physician | -2.8 (-4.5 to -1.0) |
| Shift: pharmacy only | 3.9 (2.3 to 5.5) |
| Shift: traditional or other informal | -1.2 (-2.4 to -0.1) |
| Shift: no care sought | 0.1 (-0.3 to 0.5) |

Slope index of inequality for combined treatment in each survey, without and with adjustment for characteristics (percentage points, 95% CI from the same replicate weights). The reduction was smaller in 2019 than in 2025, so the pooled reduction should not be applied to either survey alone.

| Survey and adjustment | Slope index or reduction, points (95% CI) |
| --- | --- |
| 2019: none | 10.6 (1.4 to 19.7) |
| 2019: characteristics | 9.2 (-2.2 to 20.6) |
| 2019: reduction from characteristics | 1.4 (-6.7 to 9.5) |
| 2025: none | 13.0 (3.2 to 22.8) |
| 2025: characteristics | 4.6 (-7.4 to 16.6) |
| 2025: reduction from characteristics | 8.4 (1.4 to 15.3) |
