## Supplementary material for "Oral rehydration salts and zinc for childhood diarrhoea in Bangladesh, 2019 and 2025: coverage, wealth inequality and reported source of care": S4_Table.docx

### **S4 Table. Treatment combinations, care from facilities and physicians by wealth, and where ORS and zinc were obtained**

A. Treatment combinations and where children without combined treatment were, MICS 2019 and 2025.

Treatment combinations by source of care (weighted percent of children with diarrhoea). Among children given ORS, 48.4% in 2019 and 56.6% in 2025 were also given zinc.

| Source of care | Survey | ORS and zinc | ORS only | Zinc only | Neither |
| --- | --- | --- | --- | --- | --- |
| All | 2019 | 35.0 | 37.3 | 8.6 | 19.1 |
| All | 2025 | 42.9 | 32.9 | 9.0 | 15.2 |
| Public health care | 2019 | 67.5 | 20.2 | 8.0 | 4.3 |
| Public health care | 2025 | 72.4 | 19.6 | 5.5 | 2.6 |
| Private facility or physician | 2019 | 51.7 | 26.6 | 12.6 | 9.1 |
| Private facility or physician | 2025 | 65.6 | 15.1 | 14.1 | 5.1 |
| Pharmacy only | 2019 | 36.1 | 41.1 | 9.1 | 13.7 |
| Pharmacy only | 2025 | 49.2 | 27.2 | 13.1 | 10.4 |
| Traditional or other informal | 2019 | 35.4 | 37.1 | 11.6 | 15.9 |
| Traditional or other informal | 2025 | 48.2 | 30.9 | 9.6 | 11.3 |
| No care sought | 2019 | 8.6 | 48.4 | 4.0 | 39.1 |
| No care sought | 2025 | 12.5 | 53.0 | 2.6 | 31.8 |

Share of children without combined treatment, and of children given ORS without zinc, by source of care (weighted percent, 95% CI):

| Group | Source of care | Survey | Percent (95% CI) |
| --- | --- | --- | --- |
| Not given both | Public health care | 2019 | 6.3 (4.6 to 8.0) |
| Not given both | Public health care | 2025 | 4.8 (3.2 to 6.5) |
| Not given both | Private facility or physician | 2019 | 12.6 (10.2 to 15.0) |
| Not given both | Private facility or physician | 2025 | 7.3 (5.2 to 9.4) |
| Not given both | Pharmacy only | 2019 | 25.4 (22.2 to 28.7) |
| Not given both | Pharmacy only | 2025 | 31.2 (27.8 to 34.5) |
| Not given both | Traditional or other informal | 2019 | 17.1 (14.6 to 19.6) |
| Not given both | Traditional or other informal | 2025 | 12.9 (10.3 to 15.5) |
| Not given both | No care sought | 2019 | 38.6 (35.3 to 42.0) |
| Not given both | No care sought | 2025 | 43.7 (40.0 to 47.5) |
| ORS without zinc | Public health care | 2019 | 6.8 (4.4 to 9.2) |
| ORS without zinc | Public health care | 2025 | 6.0 (3.6 to 8.4) |
| ORS without zinc | Private facility or physician | 2019 | 12.1 (9.2 to 15.0) |
| ORS without zinc | Private facility or physician | 2025 | 5.6 (3.3 to 7.9) |
| ORS without zinc | Pharmacy only | 2019 | 28.5 (24.0 to 33.0) |
| ORS without zinc | Pharmacy only | 2025 | 29.0 (24.7 to 33.4) |
| ORS without zinc | Traditional or other informal | 2019 | 17.1 (13.8 to 20.3) |
| ORS without zinc | Traditional or other informal | 2025 | 13.4 (9.9 to 16.9) |
| ORS without zinc | No care sought | 2019 | 35.6 (31.0 to 40.1) |
| ORS without zinc | No care sought | 2025 | 46.0 (41.1 to 50.9) |

Share of all children in 2025 (Table 1): public 10.0%, private facility or physician 12.2%, pharmacy-only 35.0%, traditional or other informal 14.2% and no care sought 28.5%. Share of children without combined treatment in 2025 (table above): public 4.8%, private facility or physician 7.3%, pharmacy-only 31.2%, traditional or other informal 12.9% and no care sought 43.7%. Children with no care reported are over-represented among those without combined treatment. Pharmacy-only children are under-represented, and the two groups together were 63.5% of all children.

### **B. Care from facilities and physicians by wealth, and where ORS and zinc were obtained in 2019.**

Any medical care and care from a public or private facility or physician by wealth quintile (weighted percent, 95% CI):

| Survey | Group | Any medical care | Public or private facility or physician |
| --- | --- | --- | --- |
| 2019 | Lowest (Quintile 1 / Poorest) | 48.0 (43.0 to 53.0) | 25.6 (21.1 to 30.0) |
| 2025 | Lowest (Quintile 1 / Poorest) | 51.0 (45.5 to 56.5) | 13.6 (10.1 to 17.1) |
| 2019 | Second (Quintile 2 / Poorer) | 49.7 (44.0 to 55.4) | 21.7 (16.9 to 26.5) |
| 2025 | Second (Quintile 2 / Poorer) | 52.5 (46.2 to 58.9) | 18.4 (13.7 to 23.1) |
| 2019 | Middle (Quintile 3) | 55.1 (48.3 to 61.9) | 32.7 (26.0 to 39.4) |
| 2025 | Middle (Quintile 3) | 59.9 (53.7 to 66.1) | 21.5 (16.5 to 26.5) |
| 2019 | Fourth (Quintile 4 / Richer) | 64.7 (58.2 to 71.1) | 33.0 (26.9 to 39.2) |
| 2025 | Fourth (Quintile 4 / Richer) | 59.9 (53.4 to 66.3) | 25.7 (19.9 to 31.5) |
| 2019 | Highest (Quintile 5 / Richest) | 65.6 (58.4 to 72.8) | 40.0 (32.5 to 47.5) |
| 2025 | Highest (Quintile 5 / Richest) | 64.1 (57.7 to 70.6) | 33.4 (26.8 to 40.1) |

The slope index of inequality for care from a public or private facility or physician was 20.3 points (95% CI 11.2 to 29.5) in 2019. It was 23.2 points (15.1 to 31.2) in 2025. The change between surveys was 2.8 points (-9.4 to 15.0; p=0.65).

Source of ORS and zinc among children who received them, MICS 2019 (weighted percent and standard error). When several sources were named, the first in the order public, private or NGO medical, pharmacy, other was used.

| Item | Source | Percent | SE |
| --- | --- | --- | --- |
| Zinc | Other or DK | 16.2 | 1.5 |
| Zinc | Pharmacy | 72.1 | 1.9 |
| Zinc | Private or NGO medical | 3.2 | 0.7 |
| Zinc | Public health care | 8.5 | 1.2 |
| ORS | Other or DK | 24.0 | 1.4 |
| ORS | Pharmacy | 63.9 | 1.6 |
| ORS | Private or NGO medical | 3.3 | 0.6 |
| ORS | Public health care | 8.8 | 0.9 |

Where zinc and ORS were obtained, by source of care, MICS 2019 (weighted row percent among children who received each item):

| Item | Source of care | Public health care | Private or NGO medical | Pharmacy | Other or DK |
| --- | --- | --- | --- | --- | --- |
| Zinc | Public health care | 38.5 | 0.6 | 54.6 | 6.2 |
| Zinc | Private facility or physician | 0.0 | 8.8 | 86.0 | 5.2 |
| Zinc | Pharmacy only | 0.0 | 0.0 | 98.8 | 1.2 |
| Zinc | Traditional or other informal | 0.0 | 3.4 | 36.9 | 59.7 |
| Zinc | No care sought | 1.0 | 3.3 | 68.3 | 27.5 |
| ORS | Public health care | 51.6 | 1.0 | 40.1 | 7.3 |
| ORS | Private facility or physician | 0.6 | 14.1 | 77.4 | 7.9 |
| ORS | Pharmacy only | 0.0 | 1.1 | 95.1 | 3.8 |
| ORS | Traditional or other informal | 2.8 | 0.5 | 26.1 | 70.5 |
| ORS | No care sought | 1.6 | 0.7 | 59.7 | 38.0 |

Where ORS and zinc were obtained by reported source of care, MICS 2019, as a share of all children in each group (weighted percent, 95% CI). Zinc users are children given zinc, and n is the unweighted number of children in the group. The groups defined by reported care and by place of supply overlap. Of children with no care reported, 37.7% (32.5 to 42.9) had ORS or zinc from a pharmacy, and 8.6% (5.8 to 11.4) had both products.

| Source of care | n (zinc users) | ORS from a pharmacy | Zinc from a pharmacy | ORS or zinc from a pharmacy | Zinc from a pharmacy, among zinc users |
| --- | --- | --- | --- | --- | --- |
| Public health care | 201 (145) | 35.2 (27.7 to 42.7) | 41.2 (33.5 to 48.9) | 51.1 (43.6 to 58.7) | 54.6 (45.7 to 63.6) |
| Private facility or physician | 270 (170) | 60.6 (54.1 to 67.1) | 55.3 (48.3 to 62.2) | 75.8 (70.1 to 81.5) | 86.0 (80.4 to 91.6) |
| Pharmacy only | 368 (164) | 73.5 (68.5 to 78.5) | 44.7 (38.7 to 50.6) | 84.2 (80.0 to 88.4) | 98.8 (97.2 to 100.0) |
| Traditional or other informal | 303 (140) | 19.0 (14.1 to 23.8) | 17.3 (12.5 to 22.1) | 26.6 (21.0 to 32.1) | 36.9 (27.9 to 45.9) |
| No care sought | 447 (59) | 34.0 (28.9 to 39.1) | 8.6 (5.8 to 11.4) | 37.7 (32.5 to 42.9) | 68.3 (54.8 to 81.8) |

The 68.3% of zinc users with no care reported who obtained zinc from a pharmacy rests on 59 children, with a 95% interval of 54.8 to 81.8. The upper limit for the pharmacy-only row is truncated at 100.0.
