## Supplementary material for "Oral rehydration salts and zinc for childhood diarrhoea in Bangladesh, 2019 and 2025: coverage, wealth inequality and reported source of care": S5_Table.docx

### **S5 Table. Sensitivity analyses, full models and further checks**

A. The 13 sensitivity analyses of combined ORS and zinc treatment, MICS 2019 and 2025. Values are weighted percent or slope index with 95% CI. Change is 2025 minus 2019, in percentage points, from a linear model on the pooled design. Children: unweighted number with a non-zero sampling weight. Later parts give detail where stated.

| No. | Analysis | Assumption tested | 2019 | 2025 | Change or ratio (95% CI) | Children |
| --- | --- | --- | --- | --- | --- | --- |
|  | Primary: any ORS, “don’t know” as not given |  | 35.0 (32.3 to 37.7) | 42.9 (40.1 to 45.8) | 7.9 (4.0 to 11.8) | 3,222 |
| 1 | ORS from packet only | Counting any ORS, given that the 2025 report value matches packet ORS | 34.3 (31.6 to 37.0) | 41.6 (38.8 to 44.5) | 7.3 (3.4 to 11.2) | 3,222 |
| 2 | “Don’t know” answers set to missing | “Don’t know” means not given | 35.1 (32.4 to 37.9) | 43.0 (40.2 to 45.9) | 7.9 (4.0 to 11.8) | 3,210 |
| 3 | Children aged 6 to 59 months | Including infants for whom evidence on zinc is weak | 37.2 (34.3 to 40.1) | 44.8 (41.8 to 47.8) | 7.6 (3.4 to 11.8) | 2,933 |
| 4 | Adjusted for interview month | Season does not explain the change |  |  | 8.4 (4.2 to 12.6); prevalence ratio 1.24 (1.11 to 1.39), unadjusted 1.23 | 3,222 |
| 5 | February to May only (part H) | Months sampled in only one survey do not drive the change | 34.5 (31.7 to 37.3) | 43.2 (40.3 to 46.2) | 8.7 (4.6 to 12.8) | 2,964 |
| 6 | Slope index from quintile midpoints | Reliance on individual wealth ranks | 8.9 (-0.4 to 18.3) | 12.1 (2.2 to 22.0) | 3.2 (-10.5 to 16.8); p=0.65 | 3,222 |
| 7 | Slope index, rank among all children under five | Ranking only children with diarrhoea | 10.5 (1.4 to 19.7) | 13.2 (3.6 to 22.9) | 2.7 (-10.6 to 16.0); p=0.69 | 3,222 |
| 8 | Model 3: illness proxies added (part B) | Differences in severity between sources |  |  | Pharmacy aPR 0.60 (0.53 to 0.67), against 0.59 | 3,222 |
| 9 | Infants split at six months (part E) | One group for ages 0 to 11 months |  |  | Age 0 to 5 months, aPR 0.46 (0.34 to 0.61) | 3,222 |
| 10 | Any pharmacy contact (part G) | Pharmacy-only group excludes children who also used a facility | 37.0 (31.6 to 42.4) | 50.1 (45.4 to 54.7) |  | 3,222 |
| 11 | Three other hierarchies (part F) | Order of assignment for several sources |  |  | Pharmacy aPR 0.60 to 0.61, against 0.59 | 3,222 |
| 12 | Logistic regression, marginal standardisation (part C) | Log-link Poisson model |  |  | Pharmacy ratio 0.60, against 0.59 | 3,222 |
| 13 | Private physician, pharmacy-only and traditional groups combined (part G) | The pharmacy rise is not a shift between codes | 59.9 (57.2 to 62.7) | 61.5 (58.7 to 64.2) | 1.5 (-2.4 to 5.4) | 3,222 |

B. Full models: adjusted prevalence ratios for combined treatment, pooled MICS 2019 and 2025 (aPR, 95% CI; n=3,222). Reference groups are 2019, poorest quintile, secondary education (the largest group), urban area, Dhaka division, age 12 to 23 months, boys and public care. Model 3 adds less to drink, less to eat and cough to Model 2. These are illness proxies. The drinking item asked in 2019 what the child drank and in 2025 what the child was given, so it is an imperfect proxy. No duration, stool or dehydration item exists, and fever was asked only in 2019. Adding fever to the 2019 data gave a pharmacy aPR of 0.52 and a fever aPR of 1.01 (0.87 to 1.17). With no education as the reference, the aPR in Model 1 was 0.95 (0.76 to 1.18) for primary. It was 1.09 (0.88 to 1.34) for secondary and 1.16 (0.92 to 1.47) for higher secondary or more.

| Variable | Model 1 | Model 2 | Model 3 |
| --- | --- | --- | --- |
| Survey 2025 (ref 2019) | 1.19 (1.07 to 1.32) | 1.26 (1.15 to 1.39) | 1.26 (1.14 to 1.38) |
| Wealth: Second (Quintile 2 / Poorer) | 0.90 (0.77 to 1.05) | 0.91 (0.79 to 1.04) | 0.91 (0.79 to 1.04) |
| Wealth: Middle (Quintile 3) | 0.98 (0.83 to 1.14) | 0.95 (0.82 to 1.09) | 0.94 (0.82 to 1.08) |
| Wealth: Fourth (Quintile 4 / Richer) | 1.08 (0.92 to 1.26) | 1.03 (0.89 to 1.18) | 1.03 (0.90 to 1.18) |
| Wealth: Highest (Quintile 5 / Richest) | 1.03 (0.86 to 1.24) | 0.98 (0.83 to 1.16) | 0.98 (0.82 to 1.15) |
| Education: no education | 0.92 (0.75 to 1.13) | 0.95 (0.79 to 1.13) | 0.95 (0.80 to 1.14) |
| Education: Primary | 0.87 (0.76 to 1.00) | 0.87 (0.77 to 0.99) | 0.88 (0.78 to 1.00) |
| Education: Higher secondary or more | 1.07 (0.94 to 1.22) | 1.01 (0.90 to 1.14) | 1.02 (0.91 to 1.15) |
| Rural (ref urban) | 0.98 (0.87 to 1.12) | 0.99 (0.88 to 1.11) | 0.98 (0.87 to 1.10) |
| Division: Barishal | 0.77 (0.63 to 0.93) | 0.85 (0.71 to 1.01) | 0.84 (0.71 to 1.00) |
| Division: Chattogram | 0.96 (0.83 to 1.11) | 1.04 (0.91 to 1.19) | 1.04 (0.91 to 1.19) |
| Division: Khulna | 0.92 (0.77 to 1.11) | 0.89 (0.75 to 1.05) | 0.89 (0.75 to 1.06) |
| Division: Mymensingh | 1.01 (0.80 to 1.26) | 1.04 (0.86 to 1.27) | 1.03 (0.85 to 1.26) |
| Division: Rajshahi | 0.95 (0.79 to 1.14) | 0.96 (0.81 to 1.14) | 0.98 (0.83 to 1.16) |
| Division: Rangpur | 0.83 (0.68 to 1.03) | 0.80 (0.66 to 0.97) | 0.83 (0.69 to 1.01) |
| Division: Sylhet | 0.75 (0.60 to 0.94) | 0.74 (0.60 to 0.92) | 0.74 (0.60 to 0.92) |
| Age, months: 0-11 | 0.86 (0.75 to 0.99) | 0.75 (0.67 to 0.85) | 0.78 (0.69 to 0.88) |
| Age, months: 24-35 | 1.03 (0.90 to 1.17) | 1.05 (0.93 to 1.18) | 1.05 (0.93 to 1.19) |
| Age, months: 36-47 | 1.01 (0.87 to 1.18) | 1.05 (0.92 to 1.20) | 1.07 (0.93 to 1.22) |
| Age, months: 48-59 | 0.80 (0.66 to 0.98) | 0.91 (0.76 to 1.09) | 0.92 (0.77 to 1.10) |
| Girl (ref boy) | 1.03 (0.94 to 1.14) | 1.07 (0.98 to 1.17) | 1.07 (0.98 to 1.17) |
| Care: Private facility or physician |  | 0.82 (0.72 to 0.92) | 0.83 (0.74 to 0.93) |
| Care: Pharmacy only |  | 0.59 (0.53 to 0.66) | 0.60 (0.53 to 0.67) |
| Care: Traditional or other informal |  | 0.59 (0.51 to 0.68) | 0.60 (0.52 to 0.69) |
| Care: No care sought |  | 0.14 (0.11 to 0.18) | 0.15 (0.12 to 0.18) |
| Drank or was given less to drink |  |  | 0.99 (0.90 to 1.09) |
| Ate or was given less to eat |  |  | 1.27 (1.10 to 1.46) |
| Cough in same two weeks |  |  | 0.98 (0.89 to 1.07) |

C. Checks of the source-of-care contrasts in Model 2. Year-specific aPRs come from Model 2 with a survey-by-source interaction (F(4, 1988)=1.15, p=0.33; Stata 17 gave the same result). The test on the additive scale gave p=0.10. Model 1 fitted values ranged from 0.197 to 0.591, with none above 1. In Model 2, 15 of 3,222 fitted values exceeded 1 (0.39% of the weight, maximum 1.077), all in public care in 2025. Logistic regression with marginal standardisation gives the last column.

| Source of care (ref public health care) | Model 2, pooled | 2019 | 2025 | Ratio, 2025 against 2019 | Logistic, standardised |
| --- | --- | --- | --- | --- | --- |
| Private facility or physician | 0.82 (0.72 to 0.92) | 0.76 (0.64 to 0.91) | 0.89 (0.75 to 1.04) | 1.17 (0.92 to 1.48) | 0.83 |
| Pharmacy only | 0.59 (0.53 to 0.66) | 0.51 (0.42 to 0.62) | 0.65 (0.57 to 0.76) | 1.26 (1.00 to 1.60) | 0.60 |
| Traditional or other informal | 0.59 (0.51 to 0.68) | 0.54 (0.44 to 0.66) | 0.66 (0.54 to 0.80) | 1.19 (0.90 to 1.58) | 0.60 |
| No care sought | 0.14 (0.11 to 0.18) | 0.13 (0.09 to 0.18) | 0.16 (0.12 to 0.22) | 1.31 (0.83 to 2.06) | 0.14 |

D. Exploratory tests of whether associations changed between surveys. Each row adds one survey-by-characteristic interaction to Model 1 and gives a design-based Wald test. These 18 tests were not pre-specified. The Bonferroni threshold is 0.0028, and no test passed it. Zinc coverage rose by 15.3 points in girls and 2.1 points in boys (difference 13.2, 95% CI 5.3 to 21.2), but this difference may be due to chance. These Wald tests are on the log scale. Denominator degrees of freedom are the 2,017 design degrees of freedom of the pooled subpopulation, plus one, less the number of model coefficients.

| Outcome and interaction | F (df) | p |
| --- | --- | --- |
| ORS and zinc: survey × wealth quintile | 0.85 (4) | 0.50 |
| ORS and zinc: survey × mother's education | 0.78 (3) | 0.51 |
| ORS and zinc: survey × area | 2.58 (1) | 0.11 |
| ORS and zinc: survey × division | 0.85 (7) | 0.55 |
| ORS and zinc: survey × age group | 1.13 (4) | 0.34 |
| ORS and zinc: survey × sex | 1.80 (1) | 0.18 |
| Zinc: survey × wealth quintile | 0.36 (4) | 0.84 |
| Zinc: survey × mother's education | 0.54 (3) | 0.66 |
| Zinc: survey × area | 1.22 (1) | 0.27 |
| Zinc: survey × division | 1.38 (7) | 0.21 |
| Zinc: survey × age group | 1.11 (4) | 0.35 |
| Zinc: survey × sex | 8.36 (1) | 0.004 |
| ORS: survey × wealth quintile | 1.66 (4) | 0.16 |
| ORS: survey × mother's education | 0.62 (3) | 0.60 |
| ORS: survey × area | 2.21 (1) | 0.14 |
| ORS: survey × division | 1.72 (7) | 0.099 |
| ORS: survey × age group | 1.20 (4) | 0.31 |
| ORS: survey × sex | 0.00 (1) | 0.95 |

E. Models 1 and 2 with infants split into 0 to 5 and 6 to 11 months (adjusted prevalence ratios for combined treatment, 95% CI; reference 12 to 23 months). Other covariates were as in part B (n=3,222).

| Variable | Model 1 | Model 2 |
| --- | --- | --- |
| Age, months: 0-5 | 0.46 (0.34 to 0.61) | 0.40 (0.31 to 0.53) |
| Age, months: 6-11 | 1.07 (0.94 to 1.23) | 0.94 (0.83 to 1.06) |
| Age, months: 24-35 | 1.03 (0.90 to 1.17) | 1.05 (0.93 to 1.18) |
| Age, months: 36-47 | 1.01 (0.87 to 1.18) | 1.05 (0.92 to 1.20) |
| Age, months: 48-59 | 0.80 (0.66 to 0.98) | 0.91 (0.76 to 1.08) |

F. Alternative assignment orders (hierarchies) for children who named several sources. Of care seekers, 5.7% in 2019 (60 of 1,142) and 2.6% in 2025 (36 of 1,179) named more than one type of source. The pharmacy-only group includes 18 children in 2019 and 13 in 2025 who also named an informal source. Values are pharmacy shares (percent) and aPRs from Model 2 relative to public care.

| Order of assignment | Pharmacy share, 2019 | Pharmacy share, 2025 | Private | Pharmacy | Informal | No care |
| --- | --- | --- | --- | --- | --- | --- |
| Published: public health care first | 25.9 | 35.0 | 0.82 (0.72 to 0.92) | 0.59 (0.53 to 0.66) | 0.59 (0.51 to 0.68) | 0.14 (0.11 to 0.18) |
| Pharmacy first | 27.7 | 35.8 | 0.83 (0.73 to 0.94) | 0.61 (0.54 to 0.68) | 0.59 (0.51 to 0.68) | 0.14 (0.11 to 0.18) |
| Private first | 25.9 | 35.0 | 0.84 (0.74 to 0.94) | 0.60 (0.53 to 0.67) | 0.59 (0.51 to 0.69) | 0.14 (0.11 to 0.18) |
| Informal first | 26.0 | 35.0 | 0.84 (0.74 to 0.95) | 0.61 (0.54 to 0.69) | 0.62 (0.54 to 0.71) | 0.15 (0.12 to 0.18) |

G. Further checks of wealth rank and source of care. Slope indices and changes are in percentage points. The two facility groups are public care and private facility or physician care. Any pharmacy contact includes facility users. A child’s rank is (sum of the weights of children with lower scores + half the child’s own weight) / total weight. Tied scores (79 children in 2019, 98 in 2025) were ordered as they fell in the file. The largest rank shift from using midranks was 0.0011, and midranks gave the same slope indices to two decimals. Recomputing weighted ranks within each survey in every bootstrap replicate gave the same attenuation intervals to one decimal place: 5.3 points (0.3 to 10.3) for characteristics, 5.6 (2.6 to 8.6) for care group, and 4.1 (0.7 to 7.5) for care group after characteristics. Predicted combined coverage at ranks 0 and 1 was 29.7% and 40.3% in 2019, and 36.4% and 49.4% in 2025. Squared and cubic rank terms gave p=0.81 in 2019, 0.29 in 2025 and 0.68 in the pooled data. The share of children in the combined private physician, pharmacy-only and traditional group was 59.9% (57.2 to 62.7) in 2019 and 61.5% (58.7 to 64.2) in 2025. The change was 1.5 points (-2.4 to 5.4; p=0.45). Within this group, the pharmacy-only share rose from 43.1% (39.3 to 47.0) to 57.0% (53.5 to 60.6). Combined coverage in this group was 40.3% (36.8 to 43.8) in 2019 and 52.3% (48.6 to 55.9) in 2025. Against public care, its pooled aPR was 0.64 (0.59 to 0.71). The questionnaire place codes are the same in both rounds except the NGO split (S1 Table), and interviewer manuals and Bangla wording could not be compared. A survey-by-quintile test for pharmacy-only care gave F=2.10 on 4 and 2,008 degrees of freedom (p=0.079). The facility SII changed by 2.8 points (-9.4 to 15.0; p=0.65). Combined coverage rose by 8.6 points (1.5 to 15.7) in the poorest quintile and 12.4 points (2.3 to 22.4) in the richest. The difference was 3.8 points (-8.3 to 15.9; p=0.54).

| Check | Estimate (95% CI) | p |
| --- | --- | --- |
| Zinc SII minus ORS SII, 2019 (points) | 3.0 (-9.5 to 15.5) | 0.64 |
| Zinc SII minus ORS SII, 2025 (points) | 4.8 (-6.7 to 16.2) | 0.41 |
| Combined SII, rank among all children, 2019 | 10.5 (1.4 to 19.7) |  |
| Combined SII, rank among all children, 2025 | 13.2 (3.6 to 22.9) |  |
| Change in combined SII, rank among all children | 2.7 (-10.6 to 16.0) | 0.69 |
| Any pharmacy contact, 2019 (% of children) | 27.7 (24.9 to 30.4) |  |
| Any pharmacy contact, 2025 (% of children) | 35.8 (33.1 to 38.4) |  |
| Pharmacy contact in the two facility groups, 2019 (%) | 6.2 (3.7 to 8.6) |  |
| Pharmacy contact in the two facility groups, 2025 (%) | 3.2 (1.4 to 5.0) |  |
| Combined coverage with any pharmacy contact, 2019 (%) | 37.0 (31.6 to 42.4) |  |
| Combined coverage with any pharmacy contact, 2025 (%) | 50.1 (45.4 to 54.7) |  |
| Change in SII for pharmacy-only care, 2019 to 2025 | -12.0 (-25.0 to 1.0) | 0.071 |

H. Design-based Wald tests of differences in combined coverage across each characteristic (p values). Crude tests are within each survey, with six age groups. Adjusted tests are from pooled Model 1, with five age groups.

| Characteristic | Crude, 2019 | Crude, 2025 | Adjusted, pooled |
| --- | --- | --- | --- |
| Wealth quintile | 0.096 | 0.075 | 0.27 |
| Mother’s education | 0.18 | 0.038 | 0.13 |
| Area | 0.68 | 0.033 | 0.81 |
| Division | 0.051 | 0.14 | 0.050 |
| Age group | <0.001 | 0.005 | 0.024 |
| Sex | 0.56 | 0.072 | 0.52 |
